# Evaluating the effect of a health-promoting behavioural programme on student’s quality of life, academic self-efficacy and health: Study protocol of the PROMESS-Group randomised controlled trial

**DOI:** 10.64898/2026.05.28.26354323

**Authors:** Alexandre Krikorian, Benoit Lecocq, Marina Le Pen, Aline Rollet, Evan Gouy, Mathilde Mura, Angèle Métais, Karine Spiegel, Sophie Pelloux, Julie Haesebaert, Gilles Rode, Sophie Schlatter

**Affiliations:** Faculté de Médecine Lyon Est, Université Claude Bernard, Lyon 1, Lyon, France; Research on Healthcare Performance RESHAPE, INSERM U1290, Université Claude Bernard Lyon 1, France; Research on Healthcare Performance RESHAPE, INSERM U1290, Université Claude Bernard, Lyon 1, France; Research on Healthcare Performance RESHAPE, INSERM U1290, Université Claude Bernard, Lyon 1, France, GénoPsy, Centre de Référence Maladies Rares Troubles du Comportement d’Origine Génétique (GénoPsy-Lyon), LeVinatier Psychiatrie Universitaire Lyon Métropole,Bron, France; Department of Precision Health, Luxembourg Institute of Health, Strassen, Luxembourg; Research on Healthcare Performance RESHAPE, INSERM U1290, Université Claude Bernard, Lyon 1, France, Nantes Université, Movement - Interactions - Performance, MIP, UR 4334, F-44000 Nantes, France; Universite Claude Bernard Lyon 1, CNRS, INSERM, Centre de Recherche en Neurosciences de Lyon CRNL U1028 UMR 5292, PAM Team, F-69000 Bron, France; Service de Santé Etudiante, Université Claude Bernard, Lyon 1, Lyon, France; Research on Healthcare Performance RESHAPE, INSERM U1290, Université Claude Bernard Lyon 1, France, Hospices Civils de Lyon, Pôle Santé Publique, Service Recherche et Epidémiologie Cliniques, F-69003 Lyon, France; Hospices Civils de Lyon, Unit of Physical Medicine and Rehabilitation, Henry Gabrielle Hospital, Saint-Genis-Laval, France, Univ. Lyon 1, CNRS, INSERM, Centre de Recherche en Neurosciences de Lyon CRNL U1028 UMR5292, TRAJECTOIRES, Bron, France; Research on Healthcare Performance RESHAPE, INSERM U1290, Université Claude Bernard Lyon 1, France, Claude Bernard, Lyon 1, France, Hospices Civils of Lyon, University Claude Bernard Lyon 1, Healthcare Simulation Center, SIMULYON, Lyon, France

**Keywords:** Doctoral student, health behaviour, medical student, quality of life, physical activity, sleep, stress

## Abstract

**Background:** Medical and doctoral students in health sciences represent a strategic public health lever as future professionals who will have a lasting influence on healthcare practices and the overall quality of health systems. Impaired quality of life and mental health issues among these students, coupled with scarce prevention programmes, led us to develop PROMESS-Group, an innovative multimodal programme designed to promote healthier lifestyle habits among university students.

**Methods:** We will conduct a 2-arm randomised, controlled, superiority monocentric trial to assess the effect of this programme on medical and doctoral students compared to a control group. The intervention will consist of six sessions covering stress, sleep and physical activity domains. Each session will include group and individual meetings led by trained peer experts, focusing on needs assessment, self-care education, and personalised goals setting. Students’ quality of life, academic self-efficacy, and broader health outcomes will be assessed using validated questionnaires and objective tools at baseline, during, and post-intervention. Data will be analysed according to the intention-to-treat principle and presented in accordance with CONSORT guidelines. Ethical approval was obtained from the institutional review board (IRB2025021802). All procedures will be performed in adherence to the Helsinki Declaration.

**Discussion:** This study will enable the generation of high-quality evidence to evaluate the programme’s effects on students’ quality of life and related psychosocial outcomes, and may inform evidence-based health promotion strategies in university settings.

Trial registration: ClinicalTrials.gov: NCT07030751 (https://clinicaltrials.gov/study/NCT07030751?locStr=Lyon,%20France&country=FR&state=Auvergne-Rh%C3%B4ne-Alpes&city=Lyon&cond=promess%20group&rank=1), 06.12.2025 - retrospectively registered.

This protocol study follows the SPIRIT guidelines (Appendix 1).

## Background

Medical and doctoral students are characterised by exceptionally high workloads and high expectations for performance (1–3). These students face intense social expectations and constant pressure to excel while navigating demanding work conditions and receiving limited institutional support (4–6). Although pressure in medical studies is largely driven by repetitive academic exams and demanding clinical internships (2), doctoral studies mark the transition from a student role to that of an academic professional, characterised by increasing responsibilities and autonomy (3). In addition to academic demands, doctoral students often face precarious employment conditions and uncertainty regarding their career prospects, which can be perceived as an additional psychological burden (7).

These high levels of academic demands and responsibilities drive students to overinvest in their studies at the expense of healthy lifestyle habits (11). This commitment contributes to high stress, sleep disorders, excessive sedentary behaviour, and insufficient physical activity (8–11). These factors affect their well-being, quality of life and mental health (3,12,13). Accordingly, an alarmingly high prevalence of anxiety disorders, depression and burnout has been observed in these populations (14,15). Although evidence that medical and doctoral students are more prone to physical health problems remains scarce (16), excessive sedentary behaviour and insufficient physical activity are especially concerning (8,12).

In addition to the risk of deteriorating health, poor health habits can significantly affect student learning and skill acquisition, leading to measurable declines in academic performance (3,17–20). For instance, high stress is associated with lower exam performance and increased risk of academic failure (21). Sleep deprivation and poor sleep quality impair memory and learning, reducing academic performance (22). Similarly, excessive sedentary behaviour and insufficient physical activity have been linked to lower grades (20). To assess other aspects of academic performance, studies should also consider students’ perceptions of their self-efficacy, defined as their belief in their ability to succeed in a particular situation (23). Academic self-efficacy is widely recognised as a critical predictor of performance among students (24), and students have identified it as a meaningful criterion for assessing academic success (11).

Maintaining a high quality of life and self-efficacy is vital for future healthcare and academic professionals, enabling them to maintain their health and performance during their studies and careers. A reduction in either dimension may lead to decrease the quality of professionals’ competencies, with broader implications for society. Healthcare professionals’ health status influences the quality of care they provide, and their physical, mental and emotional well-being is crucial to providing sustainable, compassionate, sensitive and effective care (25,26). Moreover, doctoral students’ well-being may be closely associated with academic productivity and study attrition risk (3,27,28). As such, medical and doctoral students in health sciences represent a strategic public health lever, as future professionals who will have a lasting influence on healthcare practices and the overall quality of health systems. Nevertheless, despite the well-documented global burden of impaired quality of life and adverse mental health outcomes among those students, preventive interventions addressing multiple health determinants simultaneously remain limited within higher education settings.

To address such challenges, universities must implement targeted and optimised interventions tailored to student needs (11). Although various programmes have been developed to manage stress, improve sleep, or promote physical activity among medical students (11,29–35), these programmes are rarely tailored to specific student constraints. Although the need for support appears equally crucial for doctoral students, no previous study has investigated such interventions in this population (36,37). The *Preventive Remediation for Optimal MEdical StudentS* (PROMESS) programme was recently developed to offer a multi-modal approach to support medical students in acquiring better health behaviour (11). The concept of intervention mapping inspired the development of PROMESS. Intervention mapping is a structured approach to developing public health interventions that actively involves relevant stakeholders throughout the process (38). The PROMESS programme - coconstructed with students and healthcare professionals - is based on three modules (Stress (39), Sleep (40) and Physical Activity (35)) and includes pedagogical content, personalised feedback and individual goals provided by more experienced peer coaches during individual meetings. The present work introduces a new version, the *Preventive Remediation for OptiMal UnivErsity StudentS Group* programme (PROMESS-Group) addressing the principal limitation (primarily time consumption) of PROMESS by providing pedagogical content during group sessions while retaining its strengths (multimodality, peer coaching and individual feedback). This work also considers diverse student profiles, including doctoral students, broadening its scope and relevance to populations with urgent needs.

The primary objective of this randomised controlled trial is to determine the effect of the PROMESS-Group programme on students’ quality of life compared to usual practices. Secondary objectives focus on students’ academic self-efficacy and broader aspects of their health (mental health, stress, sleep, physical activity, substance use and anthropometric markers). This trial also explores student satisfaction and moderators of programme efficacy. This study aims to provide solid evidence to consider the integration of multimodal health promotion programmes into university settings.

## Methods/Design

### 1. Design and settings

This randomised controlled trial with two parallel groups will be conducted at Claude Bernard University Lyon 1 (UCBL-1, Lyon, France), involving medical students from the Lyon Est and Lyon Sud faculties of medicine, and doctoral students from three doctoral schools in health sciences (NSCo, EDISS, CanBioS). The study was retrospectively registered on ClinicalTrials.gov (NCT07030751). The protocol was developed and refined through discussions with university students and health experts to enhance feasibility and relevance.

### 2. Ethics statement

The study protocol was approved by an Ethics Committee (CER-CUMG, IRB: 2025-02-18-02, Appendix 14) and complies with the ethical principles of the Declaration of Helsinki. This study will be conducted in accordance with the MR004 CNIL framework and the General Data Protection Regulation (GDPR), guaranteeing the anonymity and confidentiality of the collected data. No anticipated risks or harms were associated with participation. Given the low-risk, educational nature of the intervention, no independent steering committee or data monitoring committee was judged necessary. No specific criteria for discontinuing or modifying the intervention were predefined. Overall trial oversight and coordination are ensured by the principal investigator (S.S.). All students will receive detailed oral and written information from the principal investigator on the objectives and methods of the study and will sign an informed consent form (Appendix 2) after sufficient time for reflection prior to inclusion. Participation will be voluntary, without financial compensation. The project will be supervised by the university’s data protection officer, with GDPR compliance embedded in the protocol design and procedures. Personal data will be coded using unique participant identifiers, and identifying information will be stored separately from study data with restricted access. No formal auditing procedures are planned.

#### 3. Participants

##### 3a. Inclusion and exclusion criteria

Inclusion criteria include being at least 18 years old at the beginning of the study and enrolled in the fourth or fifth year of medical school or in a doctoral programme in health sciences at UCBL-1. No exclusion criteria were defined to allow the programme to be accessible to all students.

##### 3b. Recruitment and randomisation

Recruitment will be facilitated through both institutional and own student’s communication channels, including email invitations, lecture-hall presentations, posters, flyers and social media communication. Recruitment will continue until the target of 97 participants is reached. The recruited students will be randomised into (1:1 ratio) an intervention and a control group. Randomisation will be performed using a computer-generated randomisation sequence, stratified by study type and gender. Students in the interventional group will participate in a preventive programme called the PROMESS-Group, comprising six sessions structured around three core modules: stress, sleep and physical activity. Students allocated to the control group will follow usual practice during the intervention period and will subsequently receive a delayed and condensed version of the intervention. Participants in both groups will be allowed to continue their usual care and activities, with no restrictions on concomitant interventions. The study will run from 2025 to 2027 for the recruitment, intervention and main data collection phases, with the analysis phase planned at the end of 2027. Due to the nature of the intervention, participants and intervention providers cannot be blinded to group allocation. As such, no unblinding procedures are applicable. Outcome assessments will be performed by assessors blinded to the intervention arm and all analyses will be conducted on anonymised data.

### 4. Data collection

At the beginning of the study, students will complete a brief demographic survey, providing information on their year of study, age, gender, height, weight, smoking status, medication use and any recent health problems. During the study, students will answer a set of validated questionnaires, perform a set of fitness tests, wear an accelerometry recorder, and realize a cardiac activity recording as described below.

Data will be collected at the following time points:

- ***T_0_* Preintervention**: before the interventional period for both groups;
- ***T_1_* At intervention:** during the programme for the interventional group only;
- ***T_2_* Short-term postintervention**: two or three weeks after the programme for both groups;
- ***T_3_* Mid-term postintervention**: four months following *T_2_* for both groups.

#### 4a. Questionnaires

Information regarding score interpretation, ranges, thresholds and psychometric properties for each questionnaire is provided in Appendix 3.

##### Quality of life

At *T_0_*, *T_2_* and *T_3_*, students will answer the brief version of World Health Organization quality of life questionnaire (WHOQOL-BREF). This survey measures quality of life from a multidimensional perspective, including physical health, psychological health, social relationships and the environment (41).

##### Academic self-efficacy

At *T_0_*, *T_2_* and *T_3_*, students will answer the General Academic Self-Efficacy (GASE) scale. This scale assesses individuals’ perceptions of their overall academic self-efficacy. This scale reflects the global belief in one’s ability to plan and perform tasks required to complete an academic degree (42).

##### Psychological distress

At *T_0_*, *T_2_* and *T_3_*, students will answer the Kessler Psychological Distress Scale (KPDS) to measure nonspecific psychological distress over the past 30 days (43). In students, the scale allows for the rapid identification of moderate to severe levels of psychological distress, which are strongly associated with anxiety-depressive disorders and decreased academic functioning (44).

##### Loneliness

At *T_0_*, *T_2_* and *T_3_*, students will answer the *Échelle de Solitude de l’Université Laval* (ESUL) questionnaire (45), which is based on the Loneliness Scale (46), to assess subjective loneliness in French-speaking adults. Among university students, the ESUL is sensitive to variations in social and psychological well-being and is strongly correlated with levels of perceived anxiety, depression, and stress (47,48).

##### Substance use

At *T_0_*, *T_2_* and *T_3_*, students will answer the Alcohol, Smoking and Substance Involvement Screening Test (ASSIST-Lite) (49). This quick screening tool assesses the recent use of psychoactive substances (tobacco, alcohol, cannabis, stimulants, opioids and other substances) based on the full version of ASSIST developed by the World Health Organization (50).

##### Stress and coping strategies

At *T_0_*, students will complete the updated Social Readjustment Rating Scale (SRRS) (51,52) to quantify stressful life events experienced over the past year. The students will also complete the Big Five Inventory (BFI) questionnaire, which assesses personality traits within the five-factor model (53,54). At *T_0_*, *T_1_*, *T_2_* and *T_3_*, students will complete the Perceived Stress Scale (PSS), measuring subjective perceptions of stress in daily life (55,56). Moreover, students will complete the brief coping orientation problems experienced Inventory (BCI), assessing four coping behavioural strategies: problem solving, positive thinking, social support and avoidance (57,58).

##### Sleep and fatigue

At *T_0_*, students will complete the Composite Scale of Morningness (CSM) to determine chronotype (59,60). At *T_0_*, *T_2_* and *T_3_*, students will complete the Epworth Sleepiness Scale (ESS) to measure daytime sleepiness (61,62). At *T_0_*, *T_1_*, *T_2_* and *T_3_*, students will complete the Pittsburgh Sleep Quality Index (PSQI) to assess their level of sleep perturbances and its specific component : subjective sleep quality, sleep latency, sleep duration, habitual sleep efficiency, sleep disturbances, use of sleeping medication and daytime dysfunction (63,64). The students will also complete the Multidimensional Fatigue Inventory (MFI) at *T_0_*, *T_1_*, *T_2_* and *T_3_*, which explores the four dimensions of fatigue: general fatigue, mental fatigue, reduced activity and reduced motivation (65,66).

##### Physical activity and sedentary behaviour

At *T_0_*, *T_1_*, *T_2_* and *T_3_*, students will complete the national observatory of physical activity and sedentary behaviour (*Observatoire National de l’Activité Physique et de la Sédentarité* : ONAPS) Physical Activity Questionnaire (PAQ) to measure sedentary behaviour and physical activity during a typical week, distinguishing between domestic, professional, transport, or leisure activity situations (67). The students will also complete the Rapid Assessment of Physical Activity Questionnaire (RAPA) at *T_0_*, *T_1_*, *T_2_* and *T_3_* to document recent habits (68).

#### 4b. Accelerometry

At *T_0_* and *T_2_*, students’ sedentary behaviour, physical activity and sleep habits will be characterised using accelerometry. The students will be instructed to wear a triaxial accelerometer device with a dynamic range of ±8 g on the nondominant wrist for 21 consecutive days to capture representative patterns of daily life (GENEActiv, Activinsights, Unit 11, Harvard Industrial Estate, Kimbolton, Cambridgeshire, PE28 0NJ, United Kingdom; (69)). The students will be encouraged to wear the device 24 h/day while maintaining their usual lifestyle habits. Removal will be allowed when wearing the device could cause discomfort or injury, or in exceptional clinical circumstances. The GENEActiv device is a reliable and valid tool to measure sedentary behaviour, physical activity, rest-activity rhythms, and sleep in real-life conditions (intraclass correlation coefficient = 0.68–0.88; Kappa = 0.53; (70–73)). Data will be downloaded using GENEActiv PC software (v4.0.12) and saved in raw, binary format. The recordings will be set to a sampling frequency of 60 Hz and analysed according to GENEActiv package, GGIR R-package and Actiware Sleep Software algorithms (Actiware Sleep Software 6.3.0, Philips Respironics, Murrysville, PA, USA).

##### Sedentary behaviour and physical activity

Sedentary behaviour and physical activity intensity will be analysed using the Euclidean norm minus 1, expressed in milligravity units (mg). Standardised thresholds will be applied to classify intensity levels: inactivity (<100 mg), light physical activity (100–2020 mg), moderate activity (2020–5999 mg) and vigorous activity (≥5999 mg). The following markers will be extracted (35):

- Sedentary behaviour including daily sedentary time (min/day), number and duration of sedentary bouts and number of sedentary breaks;
- Total activity time (min/day);
- Light physical activity (LPA) and moderate-to-vigorous physical activity (MVPA) duration (min/day).

##### Sleep

To obtain a valid picture of habitual sleep behaviour researchers with previous training in sleep analysis will visually inspect each signal to check and correct automatic sleep/wake detection based on declarative information. Then, the following markers will be extracted (for more details, see (40)):

- Time in bed (h/day): Time between going to bed and getting out of bed;
- Total sleep time (h/day): Estimated amount of time spent asleep;
- Sleep efficiency (%): Ratio of total sleep time to time in bed;
- Sleep onset latency (min): Time taken to fall asleep after going to bed;
- Wake after sleep onset (min): Cumulative duration of wakefulness after initial sleep onset;
- Bedtime (hh:mm): Time when the student goes to bed and attempts to sleep;
- Sleep onset time (hh:mm): Time when the student falls asleep;
- Wake-up time (hh:mm): Time of final awakening in the morning;
- Mid-sleep point (hh:mm): Midpoint between sleep onset and wake-up times;
- Snooze time (min): Duration of the transition from wake-up to full alertness;
- Sleep regularity index: A measure of the stability of sleep–wake patterns.

As exploratory analysisis, non-parametric analyses will also be conducted to quantification of the regularity, fragmentation, timing and amplitude of 24-hour rest–activity rhythms (74).

#### 4c. Physical fitness

At *T_0_* and *T_2_*, the following set of physical tests will be performed under the supervision of a trained investigator and a physical activity instructor. Verbal encouragement will be provided to ensure optimal performance (for more details, see (35) and Appendix 4).

**Strength** (Figure 1, A). Lower limb muscle strength will be assessed using a maximal isometric knee extensor torque test. The student will be seated with hip and knee joints at 90° and arms crossed over the chest. The right upper thigh will be immobilised to the chair to minimise extraneous movement, and the ankle will be attached to a force gauge (DFS II, Chatillon Force Measurement, AMETEK STC, USA; (75,76)). The students will perform a warm-up with a 3-s intended extension at 10%, 30%, and 70% perceived maximal intensity, followed by two 5-s maximal isometric voluntary contractions. The exercises will be separated by a 1-min recovery period (77). The highest value will be used to determine the maximal isometric strength (in N) (78).

**Figure 1.**
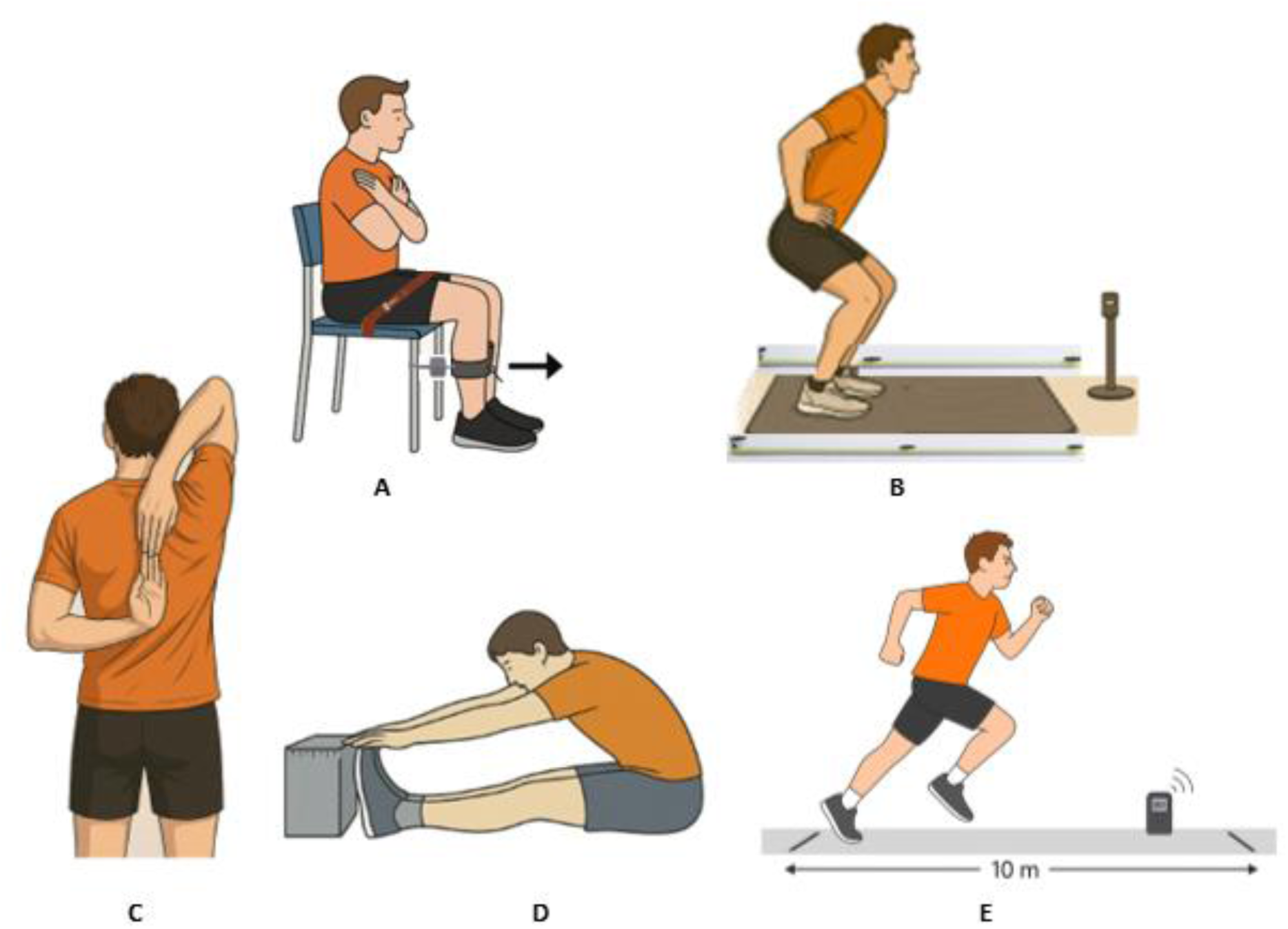
Physical fitness tests. At T_0_ (preintervention) and T_2_ (postintervention), students will perform the following physical tests under the supervision of a physical activity instructor and with verbal encouragement: A. Strength: Voluntary isometric maximal quadriceps force test. B. Power: Squat jump test. C. Flexibility: Shoulder stretch test. D. Flexibility: Sit-and-reach test. E. Endurance: 10-m shuttle run test. This Figure was generated by AI (GPT-5, OpenAI, San Francisco, CA, USA) after extensive prompting from the authors.

**Power** (Figure 1, B). The students will adopt the starting squat position and hold it for 3 s before executing a vertical jump, starting with a 90° knee angle and hands placed at the sides, performed without a previous countermovement. The students will complete two or three warm-up jumps to ensure familiarisation, followed by two jumps to reach their maximum height, with a 30-s recovery period between each. Jump heights will be recorded using the Optojump system (79). The highest jump height (cm) will be used to estimate peak power, calculated using the Sayers equation: Peak Power (W) = 60.7 × jump height (cm) + 45.3 × body mass (kg) – 2055. Relative peak power will then be obtained by dividing peak power (W) by body mass (kg) (80).

##### Flexibility

The shoulder stretch test evaluates overall shoulder mobility (Figure 1, C). The test requires the individual to place one hand behind the head and the other behind the back and try to bring the fingers together. This test can identify limitations in joint range of motion, bilateral imbalance or functional restrictions. The assessment is based on the distance between the hands (in centimetres) and may be further categorised as catching the fingers, touching them, barely touching them, not touching them or not touching the shoulder. The Sit-and-Reach Test evaluates posterior chain flexibility (Figure 1, D) (81). The test requires the individual to sit with knees fully extended and fingers aligned (one hand resting on top of the other) and perform a slow, active forward flexion of the trunk, reaching the maximal position without pain. At the end of the movement, the position is held for 2 s, during which the distance between the fingers and toes is measured.

**Endurance** (Figure 1, E). A 10-m shuttle run test will be employed to estimate cardiorespiratory fitness. The lines will be set 10 m apart, and the starting speed will be 6.5 km/h, increasing at regular intervals (+0.5 (km/h)/min) guided by an audio signal (Pack Sportbeeper Pro, CE, France). The test will stop when the individual can no longer maintain the required speed, indicating the maximal aerobic speed (*MAS*). The estimated VO_2max_ (mL/kg/min) will be calculated according to the following equation: *VO_2max_* = 8.06 * *MAS*–32.64 (35). Heart rate will be monitored during and after the run (Polar A200 device connected to a Polar H7 chest strap sensor), with recordings made immediately and at 1 and 3 min after stopping. Heart rate recovery (beats/min) will be determined by subtracting the recovery heart rate at 1 and 3 min from the heart rate at the time the individual stopped.

#### 4d. Cardiac activity

At *T_0_* and *T_2_*, cardiac activity will be recorded while students are seated in a calm environment. An additional measurement will also be performed at *T_1_* for the interventional group during the questionnaire completion period of each session. Each student will be equipped with an ear pulse sensor that will record heart rate variability (HRV) for at least 7 min (emWave Pro Plus Coherence Training software, HeartMath Institute, Boulder Creek, CA, USA). The HRV is a validated, noninvasive biomarker of autonomic nervous system regulation and reflects the dynamic balance between sympathetic and parasympathetic activity. Beyond the relevance of the HRV for stress evaluation, it is widely recognised as a marker of general health and well-being and is associated with improved cardiovascular fitness, sleep quality, emotional regulation and resilience (82–84). The following markers will be extracted via automatic analyses (35):

- RMSSD: Root Mean Square of Successive Differences in ms, quantifies short-term variations between successive heartbeats;
- SDNN: Standard Deviation of NN intervals, in which NN means normal to normal beats in ms;
- pNN50%: Percentage of number of pairs of successive intervals that differ by more than 50 ms over all NN intervals;
- HF: represents the power spectral density of activity in the 0.15– 0.40 Hz range;
- LF: represents the power spectral density of activity in the 0.04–0.15 Hz range.

#### 4.e Anthropometric markers

At *T_0_* and *T_2_*, the students’ height and weight (Scale 100, Decathlon, China) will be measured. Then, their body mass index (BMI) scores will be calculated.

### 5. Intervention

The PROMESS-Group programme is a multimodal group intervention structured around six sessions. The sessions are divided into three thematic modules: stress management, sleep improvement and physical activity promotion, each lasting from 90 to 120 min. The sessions will be led by trained peer experts (medical residents or (post)-doctoral students) and supervised by experienced facilitators. All session content is based on the PROMESS programme (refer to articles on the PROMESS Stress (39), Sleep (40), and Physical Activity (35) protocols for more information).

#### 5.a. Overall Organization

The programme will follow a coherent thematic progression, in which stress, sleep and physical activity will be progressively addressed during the six sessions (Figures 2 and 3). The order and content of the sessions was designed to follow a pedagogical and behavioural progression, considering both the students’ perceived needs and the principles of health promotion interventions. Stress, identified as the priority need, was introduced first, while physical activity, requiring greater behavioural commitment, was introduced gradually to limit the risk of early dropout. This sequence was defined by the experts based on their previous experiences. The sessions will be held every two weeks. Each session will include a group welcome, a set of questionnaires and a cardiac activity measurement (described above). The researchers will provide students with the pedagogical content based on the content of the PROMESS programme (35,39,40) and adapted for collective use. Each session will end with an individual meeting (from 20 to 30 min) during which an expert will provide personalised feedback, advice and goals to each student. During individual meetings, other students will realize module-related activities while waiting for their turn (Appendix 5).

**Figure 2:**
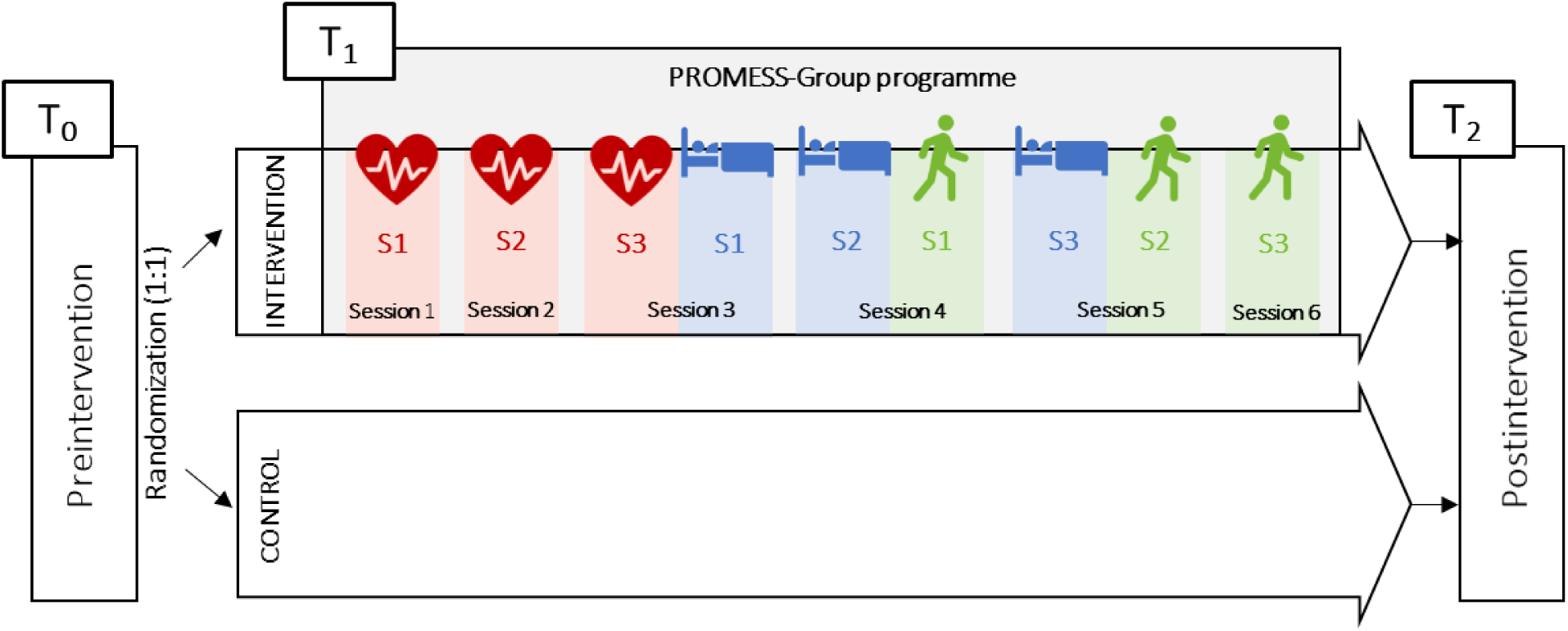
PROMESS-Group overall organisation.

**Figure 3.**
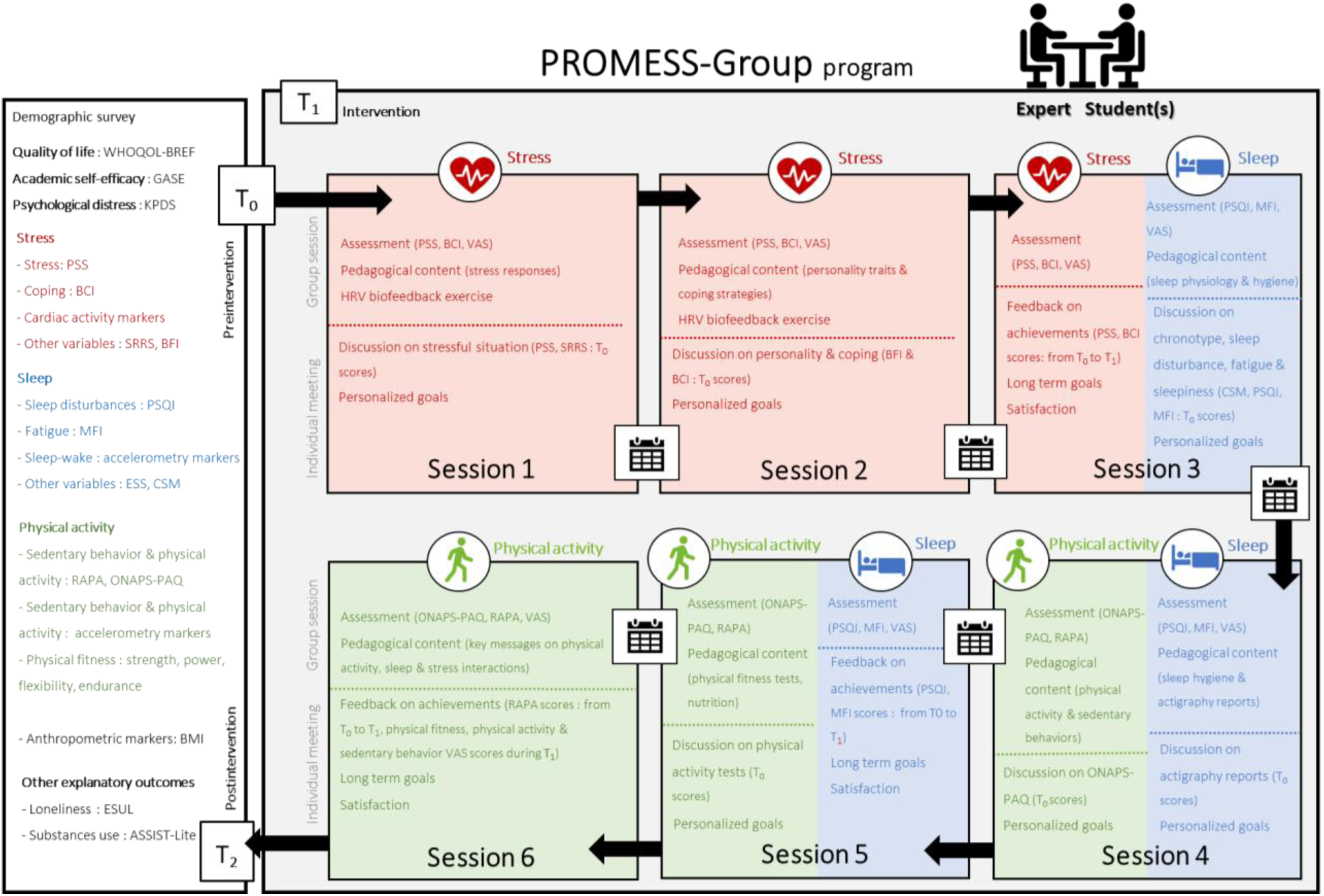
PROMESS-Group organisation for the interventional arm. At T_0_ (preintervention) and T_2_ (postintervention) students will perform a set of fitness tests, wear an accelerometry recorder, realize a cardiac activity recording, and answer a set of validated questionnaires. Questionnaires related to their quality of life (WHOQOL-BREF: World Health Organization Quality of Life: BREF questionnaire), their academic self-efficacy (GASE: General Academic Self-Efficacy), psychological distress (KPDS: Kessler Psychological Distress Scale), their stress (PSS: Perceived Stress Scale, BCI: Brief Cope Inventory, SRRS: Social Readjustment Rating Scale and BFI: Big Five Inventory), their sleep (PSQI: Pittsburgh Sleep Quality Index, MFI: Multidimensional Fatigue Inventory, ESS: Epworth Sleepiness Scale and CSM: Composite Scale of Morningness), their physical activity (RAPA: Rapid Assessment of Physical Activity and ONAPS-PAQ: ONAPS-Physical Activity Questionnaire), their loneliness (ESUL: Echelle de Solitude de l’Université Laval) and their substance use (ASSIST-Lite: Alcohol, Smoking and Substance Involvement Screening Test lite version). During the intervention (T_1_), students participate in six sessions focusing on stress, sleep and physical activity. Each session includes a group component with targeted assessments, educational content, exercises (e.g., HRV biofeedback), and an individual meeting to address difficulties and set personalised goals. The sessions will be led by trained peer experts (medical residents or (post)-doctoral students). Students will also complete a stress, sleep and physical activity diary between each session.

#### 5.b. Expert Training

All sessions will be led by experienced peers with specific training to facilitate group discussion and conduct individual meetings: They are called “experts”. The training process includes several steps: (i) reading a selection of articles (Appendix 6), (ii) becoming familiar with session procedures and observing at least one module session led by the principal investigator (S.S.) and (iii) conducting simulated individual meetings under supervision. Throughout the programme, experts will receive regular feedback from the supervisory team. This ongoing support enables qualitative monitoring and continuous improvement of practices. Systematic communication among experts also ensures consistency in delivery across sessions, contributing to the overall quality of the programme. This approach is designed to strengthen the collaborative dynamic of the programme, enhance expert engagement and foster a shared learning environment.

#### 5.c. Questionnaires

At the beginning of each session, the students will complete questionnaires and answer several 100-mm visual analogue scales (VASs; Appendix 7, 32, 36, 37). During stress sessions, students will complete the PSS and BCI questionnaires along with four VASs assessing several aspects of stress. During sleep sessions, students will answer the PSQI and MFI questionnaires, along with three VASs assessing several aspects of perceived levels of sleep and fatigue. During physical activity sessions, students will complete the ONAPS-PAQ and RAPA questionnaires and three VASs assessing several aspects of physical activity.

#### 5.d. Self-Monitoring Tools and Personalised Feedback

During each session, the experts will use consistent and interactive teaching materials. The experts will provide oral and written feedback to students, aiming to foster reflective dialogue and create a truly personalised experience. This feedback will address questionnaire responses and device-based measurements (e.g. accelerometry recordings, physical fitness tests and cardiac activity measurements) collected during the preinterventional period (*T_0_*) or during the programme (*T_1_*). In addition, students will receive a diary focused on stress, sleep and physical activity according to the ongoing modules (Appendix 8; (35,39,40). These diaries will serve as self-monitoring tools, encouraging students to observe their habits and recognise the connections between their behaviour and well-being. The diaries also promote engagement by prompting reflection on daily experiences and provide a concrete foundation for individualised feedback during following meetings. To promote the self-regulation, students will be encouraged to install two applications on their smartphones (*Apple Health* or Google Fit for daily steps and RespiRelax^+^ for cardiac coherence) and earplugs and sleep masks will be provided.

#### 5.e. Advice and Goals

During each session, students will receive practical advice and personalised goals covering stress management, fatigue reduction, sleep improvement, physical activity, fitness improvement, and reduction of sedentary behaviour. Advice and goals will be selected from a predefined list based on evidence-based recommendations and specifically designed for university students (11,35,40). Advice may include paced breathing, mindfulness, scheduling breaks, and engaging in creative, social, or physical activities (Appendix 9). Each student will commit to a small number of goals (1 to 5), selected according to their personal context, to support meaningful behavioural change. Goals will be formulated in concrete, achievable terms. To reinforce commitment, the goals will be cosigned by the expert and student (11,35,40).

#### 5.f. Detailed Session Content

All printed materials required to implement the PROMESS-Group programme are freely available from the corresponding author upon request. Figure 3 presents a content overview of each session, and Appendi× 10 provides several examples of the pedagogical content by session.

##### Session 1

This session will be devoted to stress and will begin with a group welcome session where the objectives and confidentiality principles will be presented, followed by interactive pedagogical content focusing on stress (stress cycle, physiological aspect and stress response). After contextualising the academic stressors, students will be invited to reflect on their coping strategies. Coping techniques will be briefly presented (e.g. relaxing breathing and mindfulness), focusing on HRV biofeedback techniques applicable before or after a stressful event or in regular practice (85,86,87). Then, students will perform an HRV biofeedback exercise (Appendix 11). Students will participate in an individual meeting with an expert, during which they will discuss stressful situations they encountered, their coping habits, PSS and SRRS scores (obtained at *T_0_*) and HRV biofeedback exercises. Then, the expert will set personalised goals (e.g. practising relaxing breathing for 5 min three times a day).

##### Session 2

This session will focus on gaining a deeper understanding of stress. The pedagogical content will focus on personality traits and coping strategies (problem solving, positive thinking, social support and avoidance), addressing their relationships with academic stress vulnerability and well-being (19,88,89). Then, the students will perform an HRV biofeedback exercise to estimate their resonant breathing frequency using an adapted version of the Lehrer (Appendix 11). A positive thinking strategy (i.e., positive and repetitive personal dialogue) will be introduced to help students counteract negative thoughts and facilitate emotional regulation (90). Students will participate in an individual meeting to discuss their personality (BFI) and coping scores (BCI) assessed at *T_0_*, to foster personalised reflection and support self-knowledge. The previous goals (S1) will be discussed to identify potential barriers to implementation and to adapt or set new ones.

##### Session 3

This session will conclude the stress module and initiate the sleep module. The pedagogical content will focus on the primary principles of sleep physiology, including the circadian rhythm, the sleep cycle, and the role of sleep in cognitive, emotional, and metabolic regulation. Practical recommendations will be provided on sleep hygiene, including a regular bedtime, exposure to light and an evening routine. Individual meetings will be conducted in two parts. During the first part (dedicated to stress), experts will provide students with a personalised report of the evolution of their PSS and BCI scores from *T_0_* to *T_1_* and evaluate the achievement of previously defined goals. Then, students will be invited to set their own long-term “stress management” goals. During the second part (dedicated to sleep), experts will provide information on students’ chronotypes (CSM), sleep disturbances (PSQI), fatigue (MFI) and daytime sleepiness (ESS), obtained at *T_0_*. These results will serve as the basis for discussing sleep habits, identifying primary difficulties and defining personalised goals to improve sleep and reduce fatigue (e.g. retiring at a fixed time and avoiding screens before bedtime).

##### Session 4

This session will deeply explore sleep management concepts and initiate the physical activity module. The first part will address sleep hygiene, the link between sleep and nutrition and the potential benefits of napping and sleep–wake cycles. Individual accelerometry reports will be discussed for each student (total sleep time, sleep latency, sleep efficiency and wake after onset scores, obtained at *T_0_*). The second part will introduce the physical activity module. The pedagogical content will define physical activity and sedentary behaviour, present the internationally recognised thresholds and discuss related health effects. The individual meeting will be divided into two parts. The first part (sleep) will focus on the student’s accelerometry report (*T_0_*), chronotype (*T_0_* CSM) and on previously set sleep-related goals. New, adapted goals may be established. The second part (physical activity) will address personalised feedback regarding sedentary behaviour and physical activity (ONAPS-PAQ *T_0_* scores). Then, experts will set personal goals for the students (e.g. gradually increasing the number of daily steps or reducing prolonged sedentary time by moving every 1.5 h).

##### Session 5

This session will conclude the sleep module and further explore physical activity. The session will begin with the pedagogical content on the physical fitness tests. Students will receive feedback on physical fitness test (*T_0_* scores), and specific exercises targeting relevant aspects of physical fitness will be proposed (Appendix 12). Then, a focus on critical aspects of nutrition will be introduced. The individual meeting will be in two parts. During the first part (dedicated to sleep), experts will provide students with a personalised report on the evolution of their PSQI (sleep disturbances and sleep duration) and MFI (general and mental fatigue) scores from *T_0_* to *T_1_* and evaluate the achievement of the previously defined goals. Then, students will be invited to set their own long-term “sleep” goals. During the second part (dedicated to physical activity), the experts will advise specific personalised physical exercises. Their previous goals will be discussed to identify potential barriers, and new goals will be set.

##### Session 6

This session will conclude the physical activity module and provide an overview of the entire programme. The pedagogical content will deliver critical messages concerning the associations between physical activity, sleep and nutrition and how these are related to health and cognitive functioning. During the meeting, experts will provide students with a personalised report on the evolution of their practice of moderate-to-intense physical activity (RAPA scores from *T_0_* to *T_1_*) and of their perceived levels of physical fitness, physical activity, and sedentary behaviour (VAS scores during *T_1_*). Then, experts will evaluate progress toward previously defined goals, and students will set their own long-term physical activity goals. These meetings will also include a cross-thematic discussion on stress, sleep and physical activity, reinforcing the integrative approach of the PROMESS-Group programme. The session will conclude with a reflective discussion on the knowledge and skills acquired throughout the programme. The overarching goal is to strengthen participants’ motivation and support the long-term adoption of healthy behaviour.

To minimize contamination bias, students will be asked at the end of each session not to share session content with other students, regardless of their participation in the study.

#### 5.g. Expert Perception

At the end of each session, the expert will rate the predefined advice, report all set goals and estimate whether the students have achieved the goals set from the previous session. At the end of each module, experts will report their overall satisfaction regarding the student’s progress during the stress, sleep and physical activity module, answering “Are you satisfied with the student’s progress during the module?” on a Likert scale ranging from 1 (not satisfied at all) to 5 (very satisfied; (35,39,40).

#### 5.h. Student Satisfaction

At the final session of each module, student satisfaction will be assessed using a composite satisfaction score. This score will be calculated from several module-specific 100-mm VASs (Appendix 13; (35,39,40). For each module, the satisfaction composite scores will be categorised as follows: below 30: highly negative, 30 to 44: negative, 45 to 54: neutral, 55 to 69: positive, and 70 or above: highly positive.

### 6. Control

Students in the control group will continue to follow their usual curriculum, without any interaction from the research team during the interventional period. As with all university students, they may receive additional support from institutional services (e.g., health services, sports facilities) or seek assistance outside the academic environment on their own initiative. They will complete all data collection during the scheduled initial (preintervention: *T_0_*) and final (postintervention: *T_2_* and *T_3_*) assessment periods. Between *T_2_* and *T_3_*, students in the control group will be offered a condensed version of the programme, providing all participants with access to similar educational content.

To identify any potential change during the intervention period, all students from both the control and intervention groups will be asked: ‘Do you think your participation in the PROMESS-Group project has changed your health behaviors?’ The question covers the following topics: stress, sleep, physical activity and sedentary behavior, and other behaviors not previously listed. For each topic, respondents could indicate: no change, positive change, negative change, and provide additional comments in an open-ended format.

### 7. Data management

Data will be collected at *T_0_*, *T_1_*, *T_2_* and *T_3_*, ensuring accurate longitudinal monitoring. Data management will be conducted in strict compliance with the current regulations (the GDPR and the MR004 framework of the CNIL). Personal data will be pseudonymised using unique codes assigned to each student, with the correspondence between identities and codes securely stored by the principal investigator (S.S.). Digital data will be stored on secure servers of the RESHAPE research unit, and paper documents (questionnaires and follow-up forms) will be archived in a locked room, then scanned and destroyed after processing. Only authorised members of the project team will have access to these data. The data will be retained for a regulatory period of two years after publication of the results, then destroyed in accordance with regulatory requirements.

### 8. Statistical analysis

The data analysis will be conducted after full data collection. No interim analysis is planned under normal conditions. The variables will be expressed as the mean with standard deviation and range, the median with the interquartile range or the count and percentage. The analyses will be performed according to the intention-to-treat principle, and the results will be presented in accordance with international standards (Consolidated Standards of Reporting Trials). A *p*-value < 0.05 will be considered statistically significant. All statistical model assumptions will be verified. To manage missing data, imputation will be performed to retain as much of the available data as possible. All statistical analyses will be performed using the most recent version of the R software (R Core Team, 2025; RStudio Team, 2025, v4.5.1).

#### Sample size

The sample size was calculated a priori for the primary outcome (change in composite score of quality of life between groups from T*_0_* to T*_2_*). The sample size was determined to detect a group × time interaction in a mixed linear model with random intercept for students. The variance parameters and effect size were estimated from preliminary data from a pilot study using the same model (σ²_ID = 0.315; σ²_ε = 0.162; β_interaction = 0.338), corresponding to a standardized effect of d ≈ 0.59. With a two-tailed α risk of 0.05 and 80% power, 45 students per group are required. Taking into account an expected attrition rate of 5% in the interventional group and 10% in the control group, the final size was increased to 97 students in total.

#### Outcomes

The primary outcome will be the composite quality-of-life score (WHOQOL-BREF), averaging the physical and psychological health components, directly reflecting the domains most likely to be influenced by the PROMESS-Group programme. Secondary and exploratory outcomes focus on academic and mental health and specific health-related domains (stress, sleep and physical activity) (Figure 4). To minimise errors related to multiple testing, secondary outcomes will be analysed according to a predefined hierarchical order. The study timeline, including enrolment, interventions, and assessment time points, is summarised in the SPIRIT figure (Table 1).

**Figure 4.**
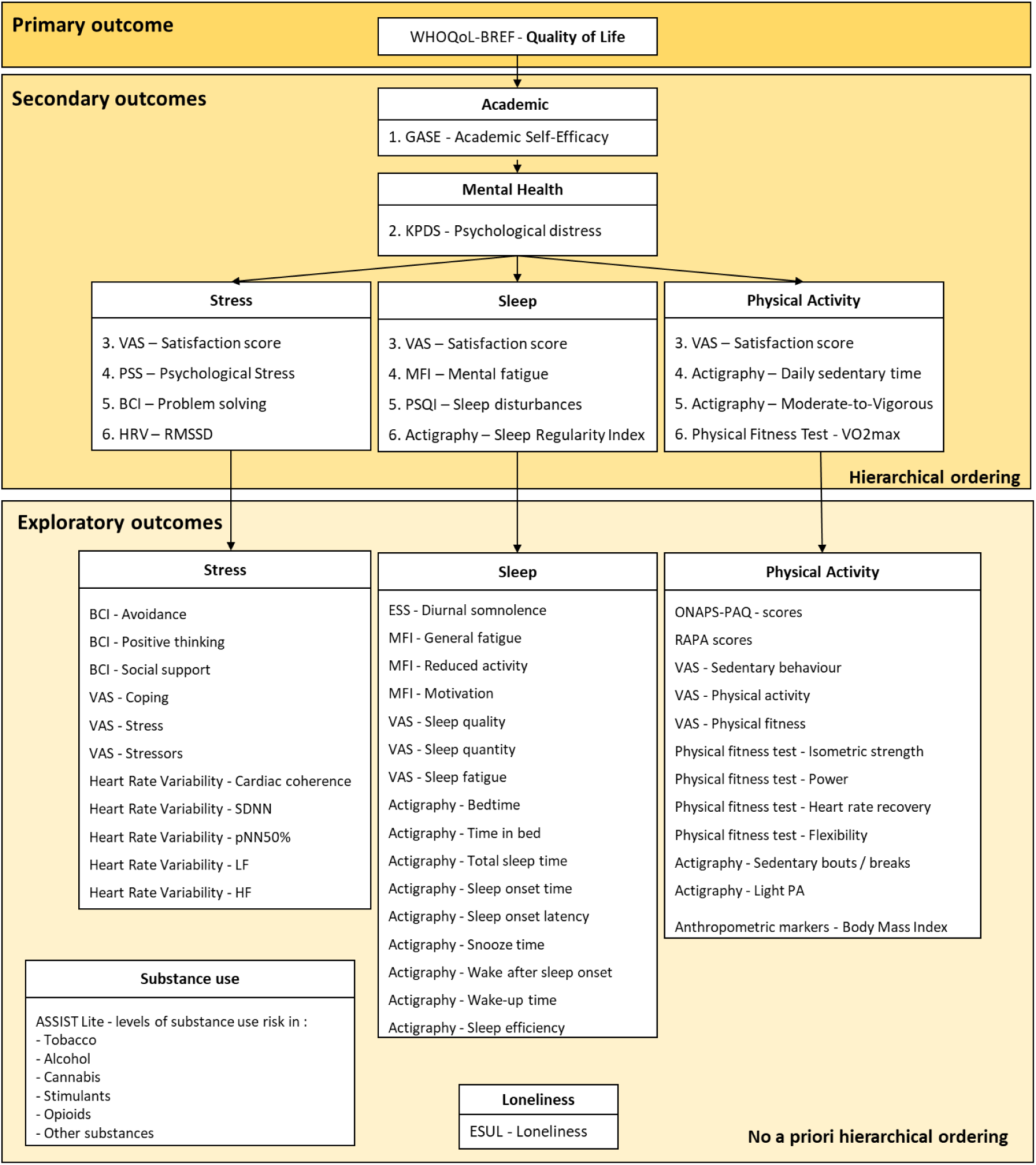
Primary, secondary and exploratory outcome classification. Abbreviations: ASSIST-Lite: Alcohol, Smoking and Substance Involvement Screening Test: Lite version, BCI: Brief COPE Inventory, BMI: Body Mass Index, ESUL: Echelle de Solitude de l’Université de Laval (Loneliness Scale), ESS: Epworth Somnolence Scale, GASE: Generalised Academic Self-Efficacy Scale, HF: high frequency, HRV: heart rate variability, LF: low frequency, MFI: Multidimensional Fatigue Inventory, PA: physical activity, pNN50%: percentage of consecutive beat-to-beat intervals that differ by more than 50ms (NN50), PSQI: Pittsburgh Sleep Quality Index, PSS: Perceived Stress Scale, RAPA: Rapid Assessment of Physical Activity, RMSSD: root mean square of successive differences, SDNN: standard deviation of NN intervals, VAS: Visual Analogue Scale, WHOQoL-BREF: brief version of the World Health Organization Quality of Life questionnaire.

**Table 1.**
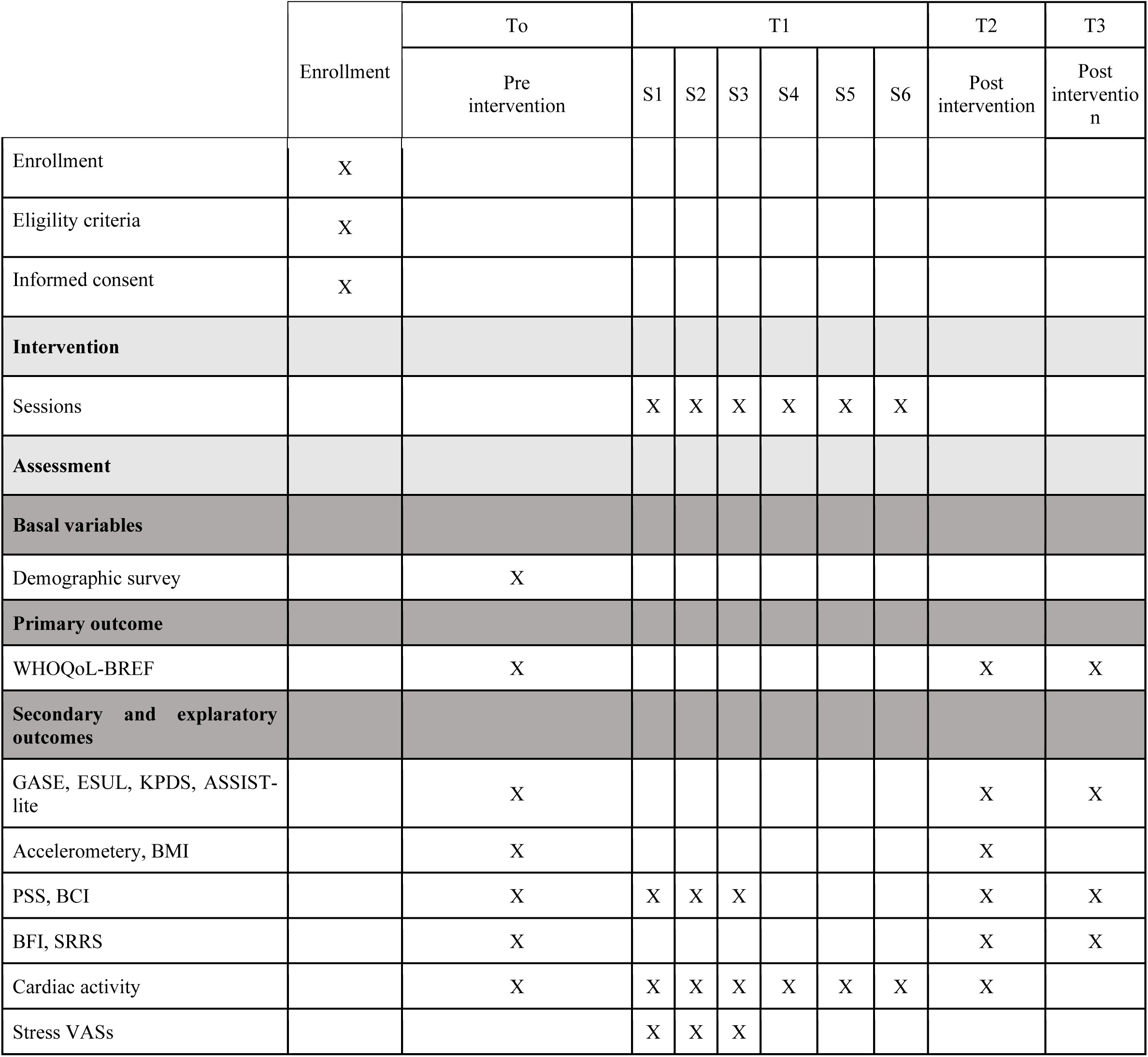
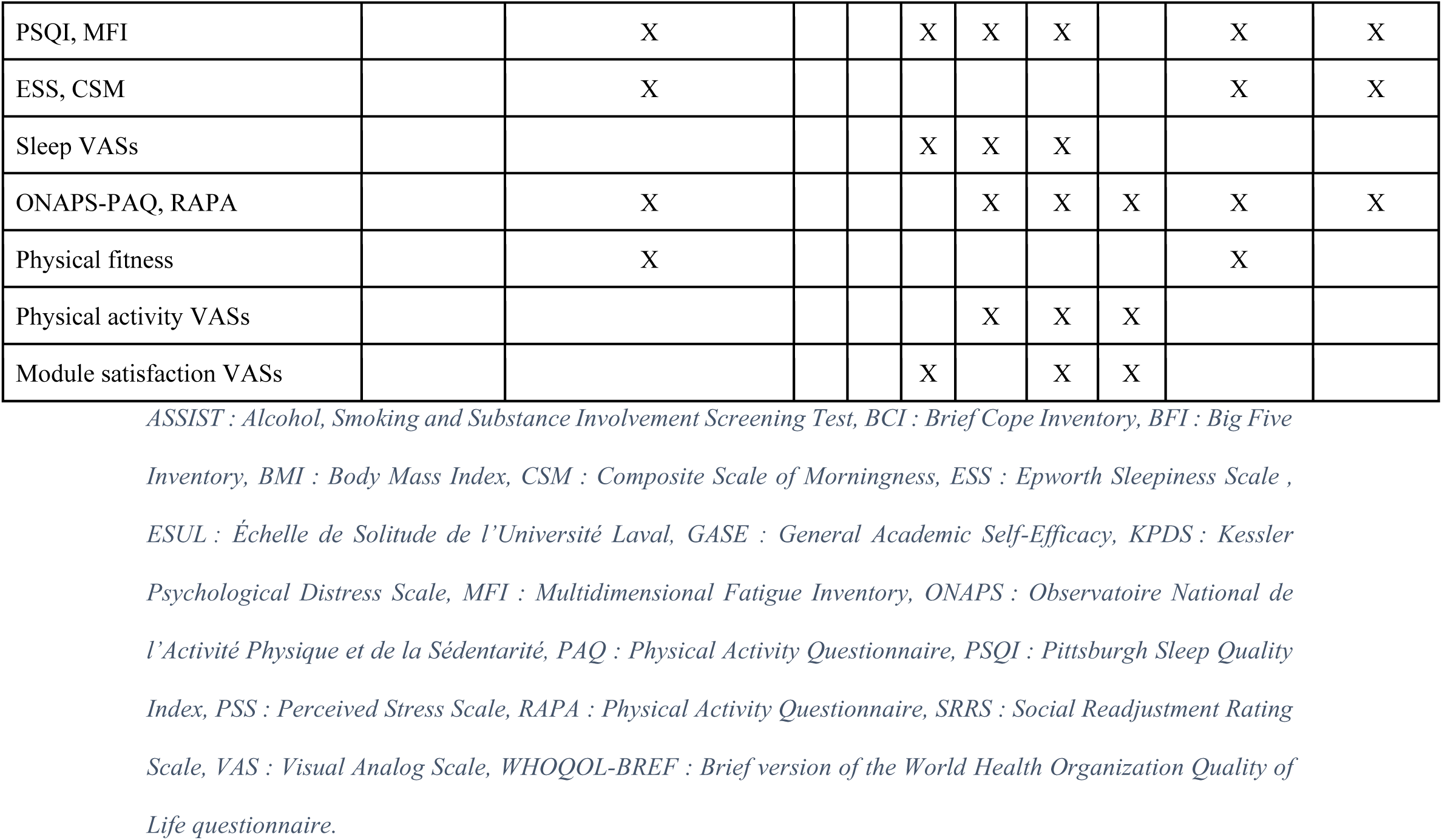
SPIRIT Figure.

#### Hierarchical ordering

An *a priori* hierarchical ordering of secondary outcomes was established (Figure 4). This ordering was defined by researchers from the PROMESS-Group, based on the existing literature, clinical relevance and expectations regarding programme efficacy. This hierarchy will guide the interpretation of the programme influence, continuing until the *p*-value becomes nonsignificant. Once a nonsignificant *p*-value is reached, the remaining secondary outcomes will be considered exploratory. For exploratory outcomes, *p*-values will be reported unadjusted and adjusted to account for multiple testing.

#### Statistical analysis plan

Linear mixed models will be employed to explore the effect of the PROMESS-Group programme on the evolution of primary, secondary and exploratory outcomes (model: outcome ∼ group*time + (1 | student)). Two types of models will be run. The first model (primary analysis) will focus on the short-term effect, with time having two categorical levels: *T_0_* (preintervention) and *T_2_* (postintervention), where no additional loss to the estimated attrition rate is expected. The second model will focus on the long-term effect, including three time levels: *T_0_*, *T_2_* and *T_3_*. Students from the control group who participate in the condensed version of the programme will be excluded. To further address potential bias introduced by this exclusion, characteristics of participants will be compared to identify potential selection bias. Moreover, a sensitivity analysis will be conducted to assess the robustness of the findings. For secondary outcomes related to satisfaction scores, the scores will be compared to 70 (i.e. the highly satisfied threshold) using either a one-sample *t*-test or a Wilcoxon signed-rank test.

#### Moderators of efficacy

The programme efficacy will be determined using the delta score on the composite quality-of-life score (*T_2_* minus *T_0_* score). A negative delta score indicates a reduced quality of life, a neutral score indicates no change, and a positive score indicates an increased quality of life during the programme. A linear model will be employed to explore whether the delta score is influenced by student characteristics at inclusion, including gender, age and status (doctoral vs medical students) and programme adherence (model: Delta ∼ gender + age + status + implementation score).

### 9. Dissemination and open science strategies

The protocol was preregistered to ensure transparency and minimise bias. Anonymous data and analysis scripts will be shared to promote reproducibility, and all materials will be available upon reasonable request to facilitate replication. The results will be disseminated via open-access publications, scientific conferences, medical thesis dissertations and doctoral student manuscripts, and will be shared with academic and educational decision-making bodies.

### 10. Use of language tools and generative artificial intelligence

Digital language assistance tools were employed to improve the editorial quality of this manuscript. A large language model (GPT-5, OpenAI, San Francisco, CA, USA) was applied on an ad hoc basis for the stylistic and linguistic rewording of some sentences. The model was also employed to generate Figure 1. A translation tool (DeepL SE, Cologne, Germany) was applied to translate some sections of the manuscript from French to English. These tools did not contribute to the scientific content or methodological design. The authors reviewed, validated and approved all final formulations of this manuscript.

## Discussion

The deterioration of the quality of life and health of medical and doctoral students represents an increasing public health concern. The PROMESS-Group protocol was designed to address these problems via a structured preventive intervention promoting a healthy lifestyle. This work is expected to enhance the students’ overall quality of life, strengthen their academic self-efficacy and better support their health. Each module is expected to yield targeted benefits: reduced stress and increased coping abilities, improved sleep quality and reduced fatigue, increased physical activity and physical fitness and decreased sedentary behaviour. Beyond the anticipated short-term effects assessed in this study, the acquisition of practical tools during the intervention may help strengthen students’ long-term self-regulation skills (e.g. stress regulation techniques, sleep hygiene routines and strategies to integrate physical activity into daily life) with possible positive implications for their long-term academic journey and future professional practice. The study results may also help inform institutional policies for prevention and health promotion in universities.

### Research implications

The PROMESS-Group programme was developed in response to students’ need for wellness support, using a comprehensive approach to ensure alignment with their expectations (11). Moreover, unlike interventions focused on a single aspect of health (30,31,33,34), the PROMESS-Group programme offers a multimodal approach that adapts well to the realities of medical and doctoral training. The ongoing study will generate a large, rich and diverse dataset, offering a unique opportunity to gain a comprehensive understanding of students’ health needs and the barriers they face during their studies. To our knowledge, this work will be the first to offer a large-scale, longitudinal assessment of the medical and doctoral student population, which is difficult to reach and evaluate. Although this programme was initially created to help medical and doctoral students, the advice and tools provided during the sessions are generalisable to those seeking to develop healthy habits and are therefore applicable to other fields. The PROMESS-Group programme is designed to be integrated into university curricula and to be reproducible and adaptable across different institutional contexts. A lighter, more educational and less research-oriented version could also be developed to facilitate large-scale implementation.

### Originality and strength

A strength of the PROMESS-Group study is the inclusion of doctoral students, a population rarely targeted by this programme type (3,8,15). Although we do not expect major differences in needs between these subgroups—since both curriculum are long and associated with heavy workloads (3,17,19) —the study will allow to test whether the programme is equally effective for both student groups. Another strength is the use of peer coaching. The programme, led by more experienced students, should benefit from enhanced legitimacy, participation and adherence (91–93). Finally, another strength is the condensed intervention for the control group, ensuring that all enrolled students can benefit from educational content, regardless of their initial group allocation. This feature promotes equity and inclusion among participants.

### Limitations

Several methodological limitations should also be mentioned. The absence of strict exclusion criteria may introduce bias. Similarly, open recruitment may bias the sample by including specific profiles (e.g. those in great need or particularly interested in this topic and/or already proficient in the behaviour). This potential selection bias could influence the generalisability of the results. However, such voluntary participation is also a prerequisite for behavioural change, as motivation and commitment are crucial elements for successful behavioural health promotion strategies. Finally, the postintervention follow-up period is set at four months. Although this window provides an interesting perspective, we cannot prove that this intervention is significantly beneficial for students in the following parts of their careers. Future work could propose a longer follow-up period and multicentre implementation.

### Conclusion

The PROMESS-Group protocol offers a multi-modal innovative approach tailored to the constraint’s students face. Its originality lies in the inclusion of doctoral students (a population rarely targeted by this programme type) and in the combination of several complementary modules. Focused on stress management, sleep improvement and physical activity promotion, the programme aims to enable action on several major health determinants that are highly prevalent in these populations. This study may open promising perspectives for the broader implementation of interventions that promote long-term well-being, health and academic success among university settings.

## Data Availability

All materials necessary to conduct the PROMESS-Group programme are available from the authors upon reasonable request. The datasets analysed during this study will be available from the corresponding author upon reasonable request. No individual participant data will be made available. In accordance with applicable data protection regulations, only aggregated and fully anonymised data will be reported in scientific publications and communications arising from this study.

## Trials Status

Version 1.0: 03/11/2026, ethical approval: 08/02/2025

Recruitment: 02/2025 to approximatively 04/2026

## List of abbreviations

AI: Artificial Intelligence
ASSIST: Alcohol, Smoking and Substance Involvement Screening Test
BCI: Brief Cope Inventory
BFI: Big Five Inventory
BMI: Body Mass Index
CER-CUMG: Comité ‘Ethique et de Recherche du Collège Universitaire de Medecine Générale
CNIL: Commission Nationale Informatique et Liberté
CSM: Composite Scale of Morningness
CVEC: Contribution de vie étudiante et de campus
EDISS: Ecole Doctorale Interdisciplinaire Sciences Santé
ESS: Epworth Sleepiness Scale
ESUL: Échelle de Solitude de l’Université Laval
GASE: General Academic Self-Efficacy
GDPR: General Data Protection Regulation
HRV: Heart Rate Variability
KPDS: Kessler Psychological Distress Scale
LPA: Light physical activity
MFI: Multidimensional Fatigue Inventory
MVPA: Moderate-to-vigorous physical activity
ONAPS: Observatoire National de l’Activité Physique et de la Sédentarité
PAQ: Physical Activity Questionnaire
PROMESS: Preventive Remediation for Optimal MEdical StudentS
PSQI: Pittsburgh Sleep Quality Index
PSS: Perceived Stress Scale
RAPA: Physical Activity Questionnaire
SRRS: Social Readjustment Rating Scale
STAPS: Sciences et Techniques des Activités Physiques et Sportives
UCBL: Université Claude Bernard Lyon 1
VAS: Visual Analog Scale
WHOQOL-BREF: Brief version of the World Health Organization Quality of Life questionnaire

## Declarations

### Ethics approval and consent to participate

This study was approved by the ethics committee of CUMG (UCBL-1) with approval number IRB 2025-02-18-02 (Appendix 14). All procedures will be performed in accordance with the Helsinki Declaration. Participants will receive oral and written information and will provide written consent (Appendix 2) before enrolment in the study after a sufficient reflection time. Participation will be voluntary, and respondents could withdraw at any time without consequence. Any important protocol amendments will be communicated to the ethics committee and updated in the trial documentation as required. No post-trial care is planned, as no risks or harms related to participation are anticipated. No compensation for harm is applicable.

### Consent for publication

All authors provided their consent to publish this article.

### SPIRIT recommendations

This manuscript follows the SPIRIT recommendation (see Table1. SPIRIT Figure and SPIRIT checklist provided in Appendix 1).

### Author contributions

**K.A.:** Conceptualisation; investigation; methodology; writing – original draught; writing – review & editing. **L.B.:** Conceptualisation; investigation; methodology; writing – original draught; writing – review & editing. **L.M.:** Conceptualisation; investigation; methodology; project administration; writing – original draught; writing – review & editing. **R.A.:** Investigation; methodology; writing – original draught; writing – review & editing. **G.E.:** Investigation; methodology; writing – review & editing. **M.M.:** Conceptualisation; investigation; methodology; writing – review & editing. **M.A.:** Conceptualisation; investigation; methodology; resources; project administration; supervision; writing – review & editing. **S.K.:** Methodology; writing – review & editing. **P.S.:** Conceptualisation; methodology; project administration; supervision; resources; writing – review & editing. **H.J.:** Conceptualisation; methodology; resources; supervision; writing – original draught; writing – review & editing. **R.G.:** Conceptualisation; resources; project administration; supervision; writing – review & editing. **S.S.:** Conceptualisation; investigation; methodology; resources; project administration; supervision; writing – original draught; writing – review & editing; contact information for trial.

## Acknowledgements

We express our gratitude to the members of the University Health Service (Université Claude Bernard Lyon 1) for their participation in the discussion and their insight into the needs of students, and their understanding of the local university system. We also thank the doctoral schools and their leaders (<u>NSCo</u>, <u>EDISS</u> , <u>CanBioS</u>) for their warm collaboration and Laura Schmidt (https://orcid.org/0000-0001-8526-5662) for her kind assistance. Last, we are grateful to the students and institutional members who believed in this project and helped secure funding (CVEC Université Claude Bernard Lyon 1).

## Author information

**A. Krikorian** is a graduate medical student at Faculté de Médecine Lyon Est, Lyon 1 Université, Lyon, France.

**B. Lecocq** is a graduate medical student at Faculté de Médecine Lyon Est, Lyon 1 Université, Lyon, France.

**M. Le Pen** is a scientific engineer at Research on Healthcare Performance RESHAPE, INSERM U1290, Lyon 1 Université, France; Faculté de Médecine Lyon Est, Université Claude Bernard Lyon 1, Lyon, France.

**A. Rollet** is a doctoral student at Research on Healthcare Performance RESHAPE, INSERM U1290, Lyon 1 Université, France; Faculté de Médecine Lyon Est, Université Claude Bernard Lyon 1, Lyon, France.

**E. Gouy** is a medical geneticist at GénoPsy (Reference Center for Rare Diseases Genetic Behavioral Disorders, Le Vinatier Psychiatrie Universitaire Lyon Métropole, teacher at Lyon 1 Université and doctoral student at Research on Healthcare Performance RESHAPE, INSERM U1290, Université Claude Bernard Lyon 1, France

**M. Mura** is a postdoctoral fellow at the Department of Precision Health, Luxembourg Institute of Health, Strassen, Luxembourg.

**A. Métais** is an associate professor, Nantes Université, Movement - Interactions - Performance, MIP, UR 4334, F-44000 Nantes, France.

**K. Spiegel** is a researcher of the French Institute of Health and Medical Research (INSERM) at Université Lyon 1 Université, CNRS, INSERM, Centre de Recherche en Neurosciences de Lyon CRNL U1028 UMR5292, PAM team, F-69500 Bron, France.

**S. Pelloux** is a general practitioner at Service de Santé Etudiante, Lyon 1 Université and the Chief Clinical Officer at the Collège Universitaire de Médecine Générale, Lyon 1 Université (Lyon, France).

**J. Haesebaert** is a professor and public health doctor at Lyon 1 Université, Lyon, France, and Université Claude Bernard Lyon 1, CNRS, INSERM, Centre de Recherche en Neurosciences de Lyon CRNL U1028 UMR5292, Trajectoires, F-69500 Bron, France.

**G. Rode** is a professor and dean at Faculté de Médecine Lyon Est, Lyon 1 Université, Lyon, France, and resaercher at the, Centre de Recherche en Neurosciences de Lyon CRNL U1028 UMR5292, CNRS, INSERM, Trajectoires, F-69500 Bron, France.

**S. Schlatter** is a postdoctoral researcher at Research on Healthcare Performance RESHAPE, INSERM U1290, Lyon 1 Université, Lyon, France.

## Supplementary information

Appendix 1. SPIRIT checklist

Appendix 2. Written consent

Appendix 3. Questionnaires

Appendix 4. Physical fitness tests

Appendix 5. Module-related activities

Appendix 6. Set of articles

Appendix 7. Visual analogue scales

Appendix 8. Stress, sleep, and physical activity diaries

Appendix 9. Lists of advices

Appendi× 10. Pedagogical content examples

Appendix 11. Cardiac coherence exercises

Appendix 12. Exercises to improve specific aspects of physical fitness

Appendix 13. Assessment of student satisfaction with modules

Appendix 14. Ethical approval

Appendix 15. Funding

## References

1. Tian-Ci Quek T, Wai-San Tam W, X. Tran B, Zhang M, Zhang Z, Su-Hui Ho C, et al. The Global Prevalence of Anxiety Among Medical Students: A Meta-Analysis. Int J Environ Res Public Health. août 2019;16(15):2735.

2. Dyrbye LN, Thomas MR, Shanafelt TD. Medical Student Distress: Causes, Consequences, and Proposed Solutions. Mayo Clin Proc. déc 2005;80(12):1613-22.

3. Schmidt M, Hansson E. Doctoral students’ well-being: a literature review. Int J Qual Stud Health Well-Being. 1 janv 2018;13(1):1508171.

4. Fares J, Al Tabosh H, Saadeddin Z, El Mouhayyar C, Aridi H. Stress, Burnout and Coping Strategies in Preclinical Medical Students. North Am J Med Sci. févr 2016;8(2):75-81.

5. Pacheco JP, Giacomin HT, Tam WW, Ribeiro TB, Arab C, Bezerra IM, et al. Mental health problems among medical students in Brazil: a systematic review and meta-analysis. Braz J Psychiatry. 31 août 2017;39(4):369-78.

6. Martinez E, Ordu C, R. Della Sala M, McFarlane A. Striving to Obtain a School-Work-Life Balance: The Full-Time Doctoral Student. Int J Dr Stud. 2013;8:039-59.

7. Huisman J, Weert E, Bartelse J. Academic Careers from a European Perspective: The Declining Desirability of the Faculty Position. J High Educ. 1 janv 2002;73:141-60.

8. Mëhilli B, Ҫabiri Z. Assessment of physical activity during PhD Studies, in Elbasan, Albania. Acad J Bus Adm Law Soc Sci. 1 juill 2025;11(2):15-24.

9. Mendelsohn D, Despot I, Gooderham PA, Singhal A, Redekop GJ, Toyota BD. Impact of work hours and sleep on well-being and burnout for physicians-in-training: the Resident Activity Tracker Evaluation Study. Med Educ. mars 2019;53(3):306-15.

10. Perotta B, Arantes-Costa FM, Enns SC, Figueiro-Filho EA, Paro H, Santos IS, et al. Sleepiness, sleep deprivation, quality of life, mental symptoms and perception of academic environment in medical students. BMC Med Educ. 17 févr 2021;21:111.

11. Métais A, Besnard L, Valero B, Henry A, Schott AM, Rode G, et al. Addressing medical students’ health challenges: codesign and pilot testing of the Preventive Remediation for Optimal MEdical StudentS (PROMESS) program. BMC Med Educ. 31 mai 2025;25(1):812.

12. Taylor CE, Scott EJ, Owen K. Physical activity, burnout and quality of life in medical students: A systematic review. Clin Teach. déc 2022;19(6):e13525.

13. Cesar FCR, Oliveira LMDAC, Ribeiro LCM, Alves AG, Moraes KL, Barbosa MA. Quality of life of master’s and doctoral students in health. Rev Bras Enferm. 2021;74(4):e20201116.

14. Rolland F, Hadouiri N, Haas-Jordache A, Gouy E, Mathieu L, Goulard A, et al. Mental health and working conditions among French medical students: A nationwide study. J Affect Disord. 1 juin 2022;306:124-30.

15. Ahalli S, Fort E, Bridai Y, Baborier N, Charbotel B. Mental health and working constraints of first-year PhD students in health and science in a French university: a cross-sectional study in the context of occupational health monitoring. BMJ Open. 30 juin 2022;12(6):e057679.

16. Hendi OM, Abdulaziz AA, Althaqafi AM, Hindi AM, Khan SA, Atalla AA. Prevalence of Musculoskeletal Disorders and its Correlation to Physical Activity Among Health Specialty Students. Int J Prev Med. 2019;10:48.

17. Seoane HA, Moschetto L, Orliacq F, Orliacq J, Serrano E, Cazenave MI, et al. Sleep disruption in medicine students and its relationship with impaired academic performance: A systematic review and meta-analysis. Sleep Med Rev. oct 2020;53:101333.

18. Stewart SM, Lam TH, Betson CL, Wong CM, Wong AM. A prospective analysis of stress and academic performance in the first two years of medical school. Med Educ. avr 1999;33(4):243-50.

19. Barret N, Guillaumée T, Rimmelé T, Cortet M, Mazza S, Duclos A, et al. Associations of coping and health-related behaviors with medical students’ well-being and performance during objective structured clinical examination. Sci Rep. 17 mai 2024;14(1):11298.

20. Chisholm-Burns MA, Berg-Poppe P, Spivey CA, Karges-Brown J, Pithan A. Systematic review of noncognitive factors influence on health professions students’ academic performance. Adv Health Sci Educ Theory Pract. oct 2021;26(4):1373-445.

21. Kötter T, Wagner J, Brüheim L, Voltmer E. Perceived Medical School stress of undergraduate medical students predicts academic performance: an observational study. BMC Med Educ. 16 déc 2017;17(1):256.

22. Phillips AJK, Clerx WM, O’Brien CS, Sano A, Barger LK, Picard RW, et al. Irregular sleep/wake patterns are associated with poorer academic performance and delayed circadian and sleep/wake timing. Sci Rep. 12 juin 2017;7(1):3216.

23. Lawrance L, McLeroy KR. Self-efficacy and health education. J Sch Health. oct 1986;56(8):317-21.

24. Hayat AA, Shateri K, Amini M, Shokrpour N. Relationships between academic self-efficacy, learning-related emotions, and metacognitive learning strategies with academic performance in medical students: a structural equation model. BMC Med Educ. déc 2020;20(1):76.

25. Fahrenkopf AM, Sectish TC, Barger LK, Sharek PJ, Lewin D, Chiang VW, et al. Rates of medication errors among depressed and burnt out residents: prospective cohort study. BMJ. 1 mars 2008;336(7642):488-91.

26. Pereira-Lima K, Mata DA, Loureiro SR, Crippa JA, Bolsoni LM, Sen S. Association Between Physician Depressive Symptoms and Medical Errors. JAMA Netw Open. 27 nov 2019;2(11):e1916097.

27. Golde CM. The Role of the Department and Discipline in Doctoral Student Attrition: Lessons from Four Departments. J High Educ. 1 nov 2005;76(6):669-700.

28. Nagy GA, Fang CM, Hish AJ, Kelly L, Nicchitta CV, Dzirasa K, et al. Burnout and Mental Health Problems in Biomedical Doctoral Students. CBE Life Sci Educ. 2019;18(2):ar27.

29. Bennett-Weston A, Keshtkar L, Jones M, Sanders C, Lewis C, Nockels K, et al. Interventions to promote medical student well-being: an overview of systematic reviews. BMJ Open. 9 mai 2024;14(5):e082910.

30. Brubaker JR, Swan A, Beverly EA. A brief intervention to reduce burnout and improve sleep quality in medical students. BMC Med Educ. 6 oct 2020;20:345.

31. Dunn LB, Iglewicz A, Moutier C. A conceptual model of medical student well-being: promoting resilience and preventing burnout. Acad Psychiatry J Am Assoc Dir Psychiatr Resid Train Assoc Acad Psychiatry. 2008;32(1):44-53.

32. Worobetz A, Retief PJ, Loughran S, Walsh J, Casey M, Hayes P, et al. A feasibility study of an exercise intervention to educate and promote health and well-being among medical students: the « MED-WELL » programme. BMC Med Educ. 3 juin 2020;20(1):183.

33. Xu YY, Wu T, Yu YJ, Li M. A randomized controlled trial of well-being therapy to promote adaptation and alleviate emotional distress among medical freshmen. BMC Med Educ. 3 juin 2019;19:182.

34. Oró P, Esquerda M, Mas B, Viñas J, Yuguero O, Pifarré J. Effectiveness of a Mindfulness-Based Programme on Perceived Stress, Psychopathological Symptomatology and Burnout in Medical Students. Mindfulness. 2021;12(5):1138-47.

35. Schlatter S, Mura M, Morel B, Loisel O, Aouidat M, Gouraud E, et al. Reducing Sedentary Behavior and Promoting Physical Activity in Medical Students: Protocol of the PROMESS-Physical Activity Clinical Trial [Internet]. Research Square; 2025 [cité 7 nov 2025]. Disponible sur: https://www.researchsquare.com/article/rs-7441836/v1

36. Solms L, Koen J, van Vianen AEM, Theeboom T, Beersma B, de Pagter APJ, et al. Simply effective? The differential effects of solution-focused and problem-focused coaching questions in a self-coaching writing exercise. Front Psychol. 18 août 2022;13:895439.

37. Williams SN, Thakore BK, McGee R. Coaching to Augment Mentoring to Achieve Faculty Diversity: A Randomized Controlled Trial. Acad Med J Assoc Am Med Coll. août 2016;91(8):1128-35.

38. Fassier JB, Lamort-Bouché M, Sarnin P, Durif-Bruckert C, Péron J, Letrilliart L, et al. [The intervention mapping protocol: A structured process to develop, implement and evaluate health promotion programs]. Rev Epidemiol Sante Publique. févr 2016;64(1):33-44.

39. Métais A, Omarjee M, Valero B, Gleich A, Mekki A, Henry A, et al. Determining the influence of an intervention of stress management on medical students’ levels of psychophysiological stress: the protocol of the PROMESS-Stress clinical trial. BMC Med Educ. 11 févr 2025;25:225.

40. Ruet A, Ndiki Mayi EF, Métais A, Valero B, Henry A, Duclos A, et al. Determining the influence of a sleep improvement intervention on medical students’ sleep and fatigue: protocol of the PROMESS-Sleep clinical trial. BMC Med Educ. 19 févr 2025;25(1):267.

41. Development of the World Health Organization WHOQOL-BREF quality of life assessment. The WHOQOL Group. Psychol Med. mai 1998;28(3):551-8.

42. Nielsen T, Dammeyer J, Vang ML, Makransky G. Gender Fairness in Self-Efficacy? A Rasch-Based Validity Study of the General Academic Self-Efficacy Scale (GASE). Scand J Educ Res. 3 sept 2018;62(5):664-81.

43. Kessler RC, Green JG, Gruber MJ, Sampson NA, Bromet E, Cuitan M, et al. Screening for serious mental illness in the general population with the K6 screening scale: results from the WHO World Mental Health (WMH) survey initiative. Int J Methods Psychiatr Res. 31 mai 2010;19(Suppl 1):4-22.

44. Stallman HM. Psychological distress in university students: A comparison with general population data. Aust Psychol. 1 déc 2010;45(4):249-57.

45. de Grâce GR, Joshi P, Pelletier R. L’Échelle de solitude de l’Université Laval (ÉSUL): validation canadienne-française du UCLA Loneliness Scale. [The Laval University loneliness scale: A Canadian-French validation of the University of California at Los Angeles (UCLA) Loneliness Scale.]. Can J Behav Sci Rev Can Sci Comport. 1993;25(1):12-27.

46. Russell D, Peplau LA, Ferguson ML. Developing a measure of loneliness. J Pers Assess. juin 1978;42(3):290-4.

47. Nicolaisen M, Thorsen K. Who are lonely? Loneliness in different age groups (18-81 years old), using two measures of loneliness. Int J Aging Hum Dev. 2014;78(3):229-57.

48. Diehl K, Jansen C, Ishchanova K, Hilger-Kolb J. Loneliness at Universities: Determinants of Emotional and Social Loneliness among Students. Int J Environ Res Public Health. 29 août 2018;15(9):1865.

49. Ali R, Meena S, Eastwood B, Richards I, Marsden J. Ultra-rapid screening for substance-use disorders: the Alcohol, Smoking and Substance Involvement Screening Test (ASSIST-Lite). Drug Alcohol Depend. 1 sept 2013;132(1-2):352-61.

50. WHO ASSIST Working Group. The Alcohol, Smoking and Substance Involvement Screening Test (ASSIST): development, reliability and feasibility. Addict Abingdon Engl. sept 2002;97(9):1183-94.

51. Holmes TH, Rahe RH. The social readjustment rating scale. J Psychosom Res. 1 août 1967;11(2):213-8.

52. Wallace D, Cooper NR, Sel A, Russo R. The social readjustment rating scale: Updated and modernised. PLOS ONE. 18 déc 2023;18(12):e0295943.

53. Plaisant O, Courtois R, Réveillère C, Mendelsohn GA, John OP. Validation par analyse factorielle du Big Five Inventory français (BFI-Fr). Analyse convergente avec le NEO-PI-R. Ann Méd-Psychol Rev Psychiatr. mars 2010;168(2):97-106.

54. John OP, Srivastava S. The Big Five Trait taxonomy: History, measurement, and theoretical perspectives. In: Handbook of personality: Theory and research, 2nd ed. New York, NY, US: Guilford Press; 1999. p. 102-38.

55. Lesage FX, Berjot S, Deschamps F. Psychometric properties of the French versions of the Perceived Stress Scale. Int J Occup Med Environ Health. juin 2012;25(2):178-84.

56. Cohen S, Kamarck T, Mermelstein R. A global measure of perceived stress. J Health Soc Behav. 1983;24(4):385-96.

57. Baumstarck K, Alessandrini M, Hamidou Z, Auquier P, Leroy T, Boyer L. Assessment of coping: a new french four-factor structure of the brief COPE inventory. Health Qual Life Outcomes. 11 janv 2017;15:8.

58. Carver CS. You want to measure coping but your protocol’s too long: consider the brief COPE. Int J Behav Med. 1997;4(1):92-100.

59. Caci H, Nadalet L, Staccini P, Myquel M, Boyer P. Psychometric properties of the French version of the composite scale of morningness in adults. Eur Psychiatry J Assoc Eur Psychiatr. sept 1999;14(5):284-90.

60. Smith CS, Reilly C, Midkiff K. Evaluation of three circadian rhythm questionnaires with suggestions for an improved measure of morningness. J Appl Psychol. oct 1989;74(5):728-38.

61. Johns MW. A New Method for Measuring Daytime Sleepiness: The Epworth Sleepiness Scale. Sleep. 1 nov 1991;14(6):540-5.

62. Kaminska M, Jobin V, Mayer P, Amyot R, Perraton-Brillon M, Bellemare F. The Epworth Sleepiness Scale: self-administration versus administration by the physician, and validation of a French version. Can Respir J. 2010;17(2):e27–34.

63. Buysse DJ, Reynolds CF, Monk TH, Berman SR, Kupfer DJ. The Pittsburgh sleep quality index: A new instrument for psychiatric practice and research. Psychiatry Res. mai 1989;28(2):193-213.

64. Blais FC, Gendron L, Mimeault V, Morin CM. [Evaluation of insomnia: validity of 3 questionnaires]. L’Encephale. 1997;23(6):447-53.

65. Gentile S, Delarozière JC, Favre F, Sambuc R, San Marco JL. Validation of the French « multidimensional fatigue inventory » (MFI 20). Eur J Cancer Care (Engl). mars 2003;12(1):58-64.

66. Smets EM, Garssen B, Bonke B, De Haes JC. The Multidimensional Fatigue Inventory (MFI) psychometric qualities of an instrument to assess fatigue. J Psychosom Res. avr 1995;39(3):315-25.

67. Charles M, Thivel D, Verney J, Isacco L, Husu P, Vähä-Ypyä H, et al. Reliability and Validity of the ONAPS Physical Activity Questionnaire in Assessing Physical Activity and Sedentary Behavior in French Adults. Int J Environ Res Public Health. 25 mai 2021;18(11):5643.

68. Topolski TD, LoGerfo J, Patrick DL, Williams B, Walwick J, Patrick MB. The Rapid Assessment of Physical Activity (RAPA) among older adults. Prev Chronic Dis. oct 2006;3(4):A118.

69. Esliger DW, Rowlands AV, Hurst TL, Catt M, Murray P, Eston RG. Validation of the GENEA Accelerometer. Med Sci Sports Exerc. juin 2011;43(6):1085-93.

70. Esliger DW, Rowlands AV, Hurst TL, Catt M, Murray P, Eston RG. Validation of the GENEA Accelerometer. Med Sci Sports Exerc. juin 2011;43(6):1085.

71. Pavey TG, Gomersall SR, Clark BK, Brown WJ. The validity of the GENEActiv wrist-worn accelerometer for measuring adult sedentary time in free living. J Sci Med Sport. mai 2016;19(5):395-9.

72. Stone JE, McGlashan EM, Facer-Childs ER, Cain SW, Phillips AJK. Accuracy of the GENEActiv Device for Measuring Light Exposure in Sleep and Circadian Research. Clocks Sleep. juin 2020;2(2):143-52.

73. Jenkins CA, Tiley LCF, Lay I, Hartmann JA, Chan JKM, Nicholas CL. Comparing GENEActiv against Actiwatch-2 over Seven Nights Using a Common Sleep Scoring Algorithm and Device-Specific Wake Thresholds. Behav Sleep Med. 2022;20(4):369-79.

74. Gonçalves BSB, Adamowicz T, Louzada FM, Moreno CR, Araujo JF. A fresh look at the use of nonparametric analysis in actimetry. Sleep Med Rev. avr 2015;20:84-91.

75. E G, P C, A GV, C F, S M, S P, et al. Impact of a submaximal mono-articular exercise on the skeletal muscle function of patients with sickle cell disease. Eur J Appl Physiol. 22 mai 2021;121(9):2459-70.

76. Mura M, Rivoire E, Dehina-Khenniche L, Weiss-Gayet M, Chazaud B, Faes C, et al. Effectiveness of an individualized home-based physical activity program in surgery-free non-endarterectomized asymptomatic stroke patients: a study protocol for the PACAPh interventional randomized trial. Trials. 14 févr 2022;23:145.

77. Petersen NT, Taylor JL, Butler JE, Gandevia SC. Depression of Activity in the Corticospinal Pathway during Human Motor Behavior after Strong Voluntary Contractions. J Neurosci. 3 sept 2003;23(22):7974-80.

78. Bachasson D, Villiot-Danger E, Verges S, Hayot M, Perez T, Chambellan A, et al. Mesure ambulatoire de la force maximale volontaire isométrique du quadriceps chez le patient BPCO. Rev Mal Respir. oct 2014;31(8):765-70.

79. Glatthorn JF, Gouge S, Nussbaumer S, Stauffacher S, Impellizzeri FM, Maffiuletti NA. Validity and reliability of Optojump photoelectric cells for estimating vertical jump height. J Strength Cond Res. févr 2011;25(2):556-60.

80. Sayers SP, Harackiewicz DV, Harman EA, Frykman PN, Rosenstein MT. Cross-validation of three jump power equations. Med Sci Sports Exerc. avr 1999;31(4):572-7.

81. Mayorga-Vega D, Merino-Marban R, Viciana J. Criterion-Related Validity of Sit-and-Reach Tests for Estimating Hamstring and Lumbar Extensibility: a Meta-Analysis. J Sports Sci Med. janv 2014;13(1):1-14.

82. Shaffer F, Meehan ZM. A Practical Guide to Resonance Frequency Assessment for Heart Rate Variability Biofeedback. Front Neurosci. 2020;14:570400.

83. Thayer JF, Åhs F, Fredrikson M, Sollers JJ, Wager TD. A meta-analysis of heart rate variability and neuroimaging studies: Implications for heart rate variability as a marker of stress and health. Neurosci Biobehav Rev. 1 févr 2012;36(2):747-56.

84. Shaffer F, Meehan ZM, Zerr CL. A Critical Review of Ultra-Short-Term Heart Rate Variability Norms Research. Front Neurosci. 19 nov 2020;14:594880.

85. Schlatter S, Schmidt L, Lilot M, Guillot A, Debarnot U. Implementing biofeedback as a proactive coping strategy: Psychological and physiological effects on anticipatory stress. Behav Res Ther. mai 2021;140:103834.

86. Schlatter ST, Thérond CC, Guillot A, Louisy SP, Duclos A, Lehot JJ, et al. Effects of relaxing breathing paired with cardiac biofeedback on performance and relaxation during critical simulated situations: a prospective randomized controlled trial. BMC Med Educ. 2 juin 2022;22(1):422.

87. Depoorter J, Hoorelbeke K, Guillaumée T, Cortet M, Lilot M, Rode G, et al. Impact of a brief HRV-biofeedback intervention on emotion regulation following a real-life stressful event: A randomized controlled study. Behaviour Research and Therapy. avr 2026;199:104979.

88. Le Saux O, Canada B, Debarnot U, Haouhache NEH, Lehot JJ, Binay M, et al. Association of Personality Traits With the Efficacy of Stress Management Interventions for Medical Students Taking Objective Structured Clinical Examinations. Acad Med. juill 2024;99(7):784-93.

89. Schlatter S, Louisy S, Canada B, Thérond C, Duclos A, Blakeley C, et al. Personality traits affect anticipatory stress vulnerability and coping effectiveness in occupational critical care situations. Sci Rep. 5 déc 2022;12(1):20965.

90. Álvarez-Pérez Y, Rivero-Santana A, Perestelo-Pérez L, Duarte-Díaz A, Ramos-García V, Toledo-Chávarri A, et al. Effectiveness of Mantra-Based Meditation on Mental Health: A Systematic Review and Meta-Analysis. Int J Environ Res Public Health. 13 mars 2022;19(6):3380.

91. Lynch TV, Beach IR, Kajtezovic S, Larkin OG, Rosen L. Step Siblings: a Novel Peer-Mentorship Program for Medical Student Wellness During USMLE Step 1 Preparation. Med Sci Educ. 13 juin 2022;32(4):803-10.

92. Snapp C, Bassett C, Baldwin A, Hill JR, DeBusk R. Peer-Assisted Learning in Undergraduate Medical Education for Resilience and Well-being. Med Sci Educ. févr 2023;33(1):5-6.

93. Abrams MP, Daly KD, Suprun A. Peer support expands wellness services and reduces mental health stigma. Med Educ. nov 2020;54(11):1050-1.

