## Additional Files for "Evaluating the effect of a health-promoting behavioural programme on student’s quality of life, academic self-efficacy and health: Study protocol of the PROMESS-Group randomised controlled trial"

### Appendix Table

Appendix 1. SPIRIT checklist 2
Appendix 2. Written consent 5
Appendix 3. Questionnaires 6
Appendix 4. Physical fitness tests 10
Appendix 5. Module-related activities 12
Appendix 6. Set of articles 13
Appendix 7. Visual analogue scales 15
Appendix 8. Stress, sleep, and physical activity diaries 16
Appendix 9. Lists of advice 19
Appendix 10. Pedagogical content examples 20
Appendix 11. Cardiac coherence exercises 26
Appendix 12. Exercises to improve specific aspects of physical fitness 27
Appendix 13. Assessment of student satisfaction with modules 28
References 29

Appendix 1. SPIRIT checklist

| Section / Topic | Item No. | SPIRIT 2025 item description | Reported on page number - Section |
|---|---|---|---|
| **Administrative information** | | | |
| Title and structured summary | 1a | Title stating trial design, population, interventions, identified as protocol | p.1 – Title |
| | 1b | Structured summary including WHO Trial Registration Data Set | p.2 – Abstract |
| Protocol version | 2 | Protocol version and date | p.6 – Methods / Design and settings |
| Roles and responsibilities | 3a | Names, affiliations, and roles of protocol contributors | p.1 – Author list; p.32 – Declarations / Author contributions |
| | 3b | Name and contact information for the trial sponsor | p.31 – Declarations / Funding |
| | 3c | Role of sponsor and funders | p.31 – Declarations / Funding |
| | 3d | Committees and oversight groups | p.7 – Methods / Ethics statement; p.31 – Declarations / Ethics approval |
| **Open science** | | | |
| Trial registration | 4 | Trial registration (registry, ID, URL, date) | p.2 – Abstract; p.6 – Methods / Design and settings |
| Protocol and statistical analysis plan | 5 | Access to protocol and statistical analysis plan | p.28 – Methods / Dissemination and open science strategies |
| Data sharing | 6 | Data sharing (IPD, code, materials) | p.28 – Methods / Dissemination and open science strategies; p.31 – Declarations / Availability of data |
| Funding and conflicts of interest | 7a | Sources of funding | p.31 – Declarations / Funding |
| | 7b | Conflicts of interest | p.31 – Declarations / Competing interests |
| Dissemination policy | 8 | Plans to communicate trial results | p.2 – Abstract / Methods; p.28 – Methods / Dissemination strategies |
| **Introduction** | | | |
| Background and rationale | 9a | Scientific background and rationale | p.4 – Background |
| | 9b | Explanation for choice of comparator | p.4 – Background; p.7 – Methods / Participants; p23 – Methods / Control |
| Objectives | 10 | Specific objectives related to benefits and harms | p.4 – Background; p7 – Methods / Ethics statement |
| **Methods: Patient and public involvement, trial design** | | | |
| Patient and public involvement | 11 | Patient and public involvement | p.6 – Methods / Design and settings |
| Trial design | 12 | Trial design (type, allocation ratio, framework) | p.6 – Methods |
| **Methods: Participants, interventions, outcomes** | | | |
| Trial setting | 13 | Trial setting | p.6 – Methods |
| Eligibility criteria | 14a | Eligibility criteria for participants | p.7 – Methods / Participants |
| | 14b | Eligibility criteria for sites and intervention providers | p.16 – Methods / Expert training |
| Intervention and comparator | 15a | Intervention and comparator description | p.15 – Methods / Intervention; p.23 – Methods / Control |
| | 15b | Criteria for modifying/discontinuing intervention | p.7 – Ethics statement |
| | 15c | Strategies to improve adherence | p.17 – Methods / Self-monitoring tools and personalized feedback |

| Section / Topic | Item No. | SPIRIT 2025 item description | Reported on page number - Section |
|---|---|---|---|
| | 15d | Concomitant care | p.7 – Methods / Participants |
| Outcomes | 16 | Primary and secondary outcomes (metrics, time points) | p.25 – Methods / Outcomes; Figure 4 |
| Harms | 17 | Harms definition and assessment | p.7 – Methods / Ethics statement |
| Participant timeline | 18 | Time schedule of enrollment, interventions (including any run-ins and washouts), assessments, and visits for participants. | p.8 – Methods / Data collection; Figures 2 & 3 |
| Sample size | 19 | Sample size calculation | p.24 – Methods / Sample size |
| Recruitment | 20 | Recruitment strategies | p.7 – Methods / Participants |
| **Methods: Assignment of interventions** | | | |
| Sequence generation | 21a | Sequence generation | p.7 – Methods / Participants |
| | 21b | Type of randomisation and stratification | p.7 – Methods / Participants |
| Allocation concealment mechanism | 22 | Allocation concealment mechanism | p.7 – Methods / Participants |
| Implementation | 23 | Implementation (access to allocation sequence) | p.7 – Methods / Participants |
| Blinding | 24a | Blinding | p.7 – Methods / Participants |
| | 24b | Blinding procedure | p.7 – Methods / Participants |
| | 24c | Unblinding | p.7 – Methods / Participants |
| **Methods: Data collection, management, analysis** | | | |
| Data collection methods | 25a | Data collection methods and instruments | p.8 – Methods / Data collection |
| | 25b | Participant retention and follow-up | p.8 – Methods / Data collection |
| Data management | 26 | Plans for data entry, coding, security, and storage | p.23 – Methods / Data management |
| Statistical methods | 27a | Statistical methods | p.24 – Methods / Statistical analysis |
| | 27b | Analysis population | p.24 – Methods / Statistical analysis |
| | 27c | Missing data handling | p.24 – Methods / Statistical analysis |
| | 27d | Additional analyses | p.27 – Methods / Statistical analysis |
| **Methods: Monitoring** | | | |
| Data monitoring committee | 28a | Data monitoring committee | p.24 – Methods / Statistical analysis |
| | 28b | Interim analyses and stopping rules | p.7 – Methods / Ethics statement |
| Trail monitoring | 29 | Frequency and procedures for monitoring trial conduct | p.7 – Methods / Ethics statement |
| **Ethics** | | | |
| Research ethics approval | 30 | Research ethics approval | p.31 – Declarations / Ethics approval and consent to participate |
| Protocol amendments | 31 | Protocol amendments | p.31 – Declarations / Ethics approval and consent to participate |
| Consent or assent | 32a | Informed consent | p.7 – Methods / Ethics statement |
| | 32b | Additional consent for ancillary studies | p.7 – Methods / Ethics statement |
| Confidentiality | 33 | How personal information about potential and enrolled participants will be collected, shared, and maintained in order to protect confidentiality before, during, and after the trial | p.7 – Methods / Ethics statement; p.23 – Methods / Data management |
| Ancillary and post-trial care | 34 | Provisions, if any, for ancillary and post-trial care, and for compensation to those who suffer harm from trial participation | p.31 – Declarations / Ethics approval and consent to participate |

| | Enrollment | To | T1 | | | | | | T2 | T3 |
|---|---|---|---|---|---|---|---|---|---|---|
| | | Pre intervention | S1 | S2 | S3 | S4 | S5 | S6 | Post intervention | Post intervention |
| **Enrollment** | X | | | | | | | | | |
| **Eligility criteria** | X | | | | | | | | | |
| **Informed consent** | X | | | | | | | | | |
| **Intervention** | | | | | | | | | | |
| **Sessions** | | | X | X | X | X | X | X | | |
| **Assessment** | | | | | | | | | | |
| **Basal variables** | | | | | | | | | | |
| **Demographic survey** | | X | | | | | | | | |
| **Primary outcome** | | | | | | | | | | |
| **WHOQoL-BREF** | | X | | | | | | | X | X |
| **Secondary and explaratory outcomes** | | | | | | | | | | |
| **GASE, ESUL, KPDS, ASSIST-lite** | | X | | | | | | | X | X |
| **Accelerometery, BMI** | | X | | | | | | | X | |
| **PSS, BCI** | | X | X | X | X | | | | X | X |
| **BFI, SRRS** | | X | | | | | | | X | X |
| **Cardiac activity** | | X | X | X | X | X | X | X | X | |
| **Stress VASs** | | | X | X | X | | | | | |

| | | | | | | | | | | |
|---|---|---|---|---|---|---|---|---|---|---|
| **PSQI, MFI** | | **X** | | | **X** | **X** | **X** | | **X** | **X** |
| **ESS, CSM** | | **X** | | | | | | | **X** | **X** |
| **Sleep VASs** | | | | | **X** | **X** | **X** | | | |
| **ONAPS-PAQ, RAPA** | | **X** | | | | **X** | **X** | **X** | **X** | **X** |
| **Physical fitness** | | **X** | | | | | | | **X** | |
| **Physical activity VASs** | | | | | | **X** | **X** | **X** | | |
| **Module satisfaction VASs** | | | | | **X** | | **X** | **X** | | |

**ASSIST** : Alcohol, Smoking and Substance Involvement Screening Test
**BCI** : Brief Cope Inventory
**BFI** : Big Five Inventory
**BMI** : Body Mass Index
**CSM** : Composite Scale of Morningness
**ESS** : Epworth Sleepiness Scale
**ESUL** : Échelle de Solitude de l'Université Laval
**GASE** : General Academic Self-Efficacy
**KPDS** : Kessler Psychological Distress Scale
**MFI** : Multidimensional Fatigue Inventory
**ONAPS** : Observatoire National de l'Activité Physique et de la Sédentarité
**PAQ** : Physical Activity Questionnaire
**PSQI** : Pittsburgh Sleep Quality Index
**PSS** : Perceived Stress Scale
**RAPA** : Physical Activity Questionnaire
**SRRS** : Social Readjustment Rating Scale
**VAS** : Visual Analog Scale
**WHOQOL-BREF** : Brief version of the World Health Organization Quality of Life questionnaire

#### Appendix 2. Written consent

**COMITÉ D'ÉTHIQUE DE LA RECHERCHE**

**FORMULAIRE DE CONSENTEMENT**

**POUR PARTICIPATION À UNE RECHERCHE**

**TITRE DE LA RECHERCHE :** Intérêt d'une intervention groupale et multimodale (*Preventive Remediation for OptiMal Students* PROMESS-Group : remédiation du stress, amélioration du sommeil et promotion de l'activité physique) sur la qualité de vie des étudiants en santé: étude interventionnelle randomisée contrôlée monocentrique.

---

Je soussigné.e ………………………………………………………… *(nom et prénom du sujet),* accepte de participer à l'étude PROMESS-Group *Preventive Remediation for OptiMal Students* : remédiation du stress, amélioration du sommeil et promotion de l'activité physique) – Intérêt d'une intervention groupale et multimodale sur la qualité de vie des étudiants en santé : étude interventionnelle randomisée contrôlée monocentrique.

Les objectifs et modalités de l'étude m'ont été clairement expliqués par le Dr Sophie SCHLATTER et/ou par la note d'information. J'ai lu et compris la fiche d'information qui m'a été remise.

À l'exception des responsables de l'étude, qui traiteront les informations dans le plus strict respect du secret médical, mon anonymat sera préservé.

J'accepte que les données nominatives me concernant recueillies à l'occasion de cette étude puissent faire l'objet d'un traitement automatisé par les organisateurs de la recherche. Je pourrai exercer mon droit d'accès et de rectification auprès du Dr Sophie SCHLATTER.

J'ai bien compris que ma participation à l'étude est volontaire.

Je suis libre d'accepter ou de refuser de participer, et je suis libre d'arrêter à tout moment ma participation en cours d'étude. Dans ce dernier cas, je contacte la responsable de l'étude, le Dr Sophie SCHLATTER, tél : +33 7 68 21 05 51,.

Mon consentement ne décharge pas les organisateurs de cette étude de leurs responsabilités. Je conserve tous mes droits garantis par la loi.

Après en avoir éventuellement discuté et avoir obtenu la réponse à toutes mes questions, j'accepte librement et volontairement de participer à la recherche qui m'est proposée.

Je m'engage à suivre l'ensemble des séances avec assiduité.

*Nom et signature de l'investigateur* *Nom et signature du sujet*

*SOPHIE SCHLATTER* *Fait à ………………… le ..../…./………*

Appendix 3. Questionnaires

| **Domain Questionnaire Item** | **Questionnaire source** | **Dimension** | **Min-max - Thresholds** | **Interpretation** | **English validation Internal consistency (Cronbach's α)** | | **French validation Internal consistency (Cronbach's α)** | |
|---|---|---|---|---|---|---|---|---|
| Quality of life<br>**WHOQOL-BREF questionnaire**<br>26 items | Development of the World Health Organization WHOQOL-BREF quality of life assessment. The WHOQOL Group. Psychol Med. mai 1998;28(3):551-8 | Physical health | 0–100 | A higher score indicates better perceived physical health. | Development of the World Health Organization WHOQOL-BREF quality of life assessment. The WHOQOL Group. Psychol Med. mai 1998;28(3):551-8 | 0.80 | Baumann C, Erpelding ML, Régat S, Collin JF, Briançon S. The WHOQOL-BREF questionnaire: French adult population norms for the physical health, psychological health and social relationship dimensions. Rev Epidemiol Sante Publique. févr 2010;58(1):33-9 | 0.74 |
| | | Psychological health | 0–100 | A higher score indicates better perceived psychological health. | | 0.76 | | 0.59 |
| | | Social relationships | 0–100 | A higher score indicates better perceived social relationships. | | 0.66 | | 0.62 |
| | | Environment | 0–100 | A higher score indicates a better perceived quality of the environment. | | 0.80 | / | / |
| Academic self-efficacy<br>**General Academic Self-Efficacy Scale**<br>5 items | Nielsen, T., Dammeyer, J., Vang, M. L., & Makransky, G. (2017). Gender fairness in self-efficacy? A Rasch-based validity study of the General Academic Self-Efficacy Scale (GASE). Scandinavian Journal of Educational Research, 62(5), 664–681 | / | 5–20 | A higher score indicates better perceived academic self-efficacy. | van Zyl, L. E., Klibert, J., Shankland, R., See-To, E. W. K., & Rothmann, S. (2022). The General Academic Self-Efficacy Scale: Psychometric properties, longitudinal invariance, and criterion validity. Journal of Psychoeducational Assessment, 40(6), 777-789. | 0.74 – 0.78 | / | / |

| | | | | | | | | |
|---|---|---|---|---|---|---|---|---|
| Psychological distress<br><br>**Kessler Psychological Distress Scale**<br><br>6 items | Kessler, R. C., Barker, P. R., Colpe, L. J., Epstein, J. F., Gfroerer, J. C., Hiripi, E., Zaslavsky, A. M. (2003). Screening for serious mental illness in the general population. Archives of General Psychiatry, 60(2), 184-189 | / | 0–24<br>-<br>≥ 13: high probability of serious mental illness | A higher score indicates a greater likelihood of having a mental disorder. | Kessler, R. C., Barker, P. R., Colpe, L. J., Epstein, J. F., Gfroerer, J. C., Hiripi, E., Zaslavsky, A. M. (2003). Screening for serious mental illness in the general population. Archives of General Psychiatry, 60(2), 184-189 | 0.89 | Arnaud B, Malet L, Teissedre F, Izaute M, Moustafa F, Geneste J, Schmidt J, Llorca PM, Brousse G. Validity study of Kessler's psychological distress scales conducted among patients admitted to French emergency department for alcohol consumption-related disorders. Alcohol Clin Exp Res. 2010 Jul;34(7):1235-45. doi: 10.1111/j.1530-0277.2010.01201.x. Epub 2010 May 7. PMID: 20477768. | 0.76 |
| Loneliness<br><br>**Échelle de Solitude de l'Université Laval**<br><br>20 items | Russell D, Peplau LA, Ferguson ML. Developing a measure of loneliness. J Pers Assess. 1978 Jun;42(3):290-4. doi: 10.1207/s15327752jpa4203_11. PMID: 660402 | / | 20–80<br>-<br>20–34: low<br>35–49: moderate<br>50–64: high | A higher score indicates higher perceived loneliness. | Russell, D., Peplau, L.A., et Cutrona, C.E. (1980). The revised UCLA Loneliness Scale. Journal of Personality and Social Psychology, 39(3), 472-480 | 0.94 | de Grâce, G.-R., Joshi, P., & Pelletier, R. (1993). L'Échelle de solitude de l'Université Laval (ÉSUL): validation canadienne-française du UCLA Loneliness Scale [The Laval University loneliness scale: A Canadian-French validation of the University of California at Los Angeles (UCLA) Loneliness Scale]. Canadian Journal of Behavioural Science/Revue canadienne des sciences du comportement, 25(1), 12–27 | 0.88 |
| Substance use<br><br>**Alcohol, Smoking and Substance Involvement Screening Test – Lite**<br><br>6 items | Ali R, Meena S, Eastwood B, Richards I, Marsden J. Ultra-rapid screening for substance-use disorders: the Alcohol, Smoking and Substance Involvement Screening Test (ASSIST-Lite). Drug Alcohol Depend. 2013 Sep 1;132(1-2):352-61. | Alcohol | 0–4<br>-<br>0–1: low<br>2: moderate<br>3–4: high | A higher score indicates higher use. | Ali R, Meena S, Eastwood B, Richards I, Marsden J. Ultra-rapid screening for substance-use disorders: The Alcohol, Smoking and Substance Involvement Screening Test (ASSIST-Lite). Drug Alcohol Depend. 2013 Sep 1;132(1- | / | / | / |
| | | Tobacco | 0–3<br>-<br>0: low<br>1–2: moderate | | | / | / | / |
| | | Cannabis | | | | / | / | / |
| | | Stimulants | | | | / | / | / |
| | | Sedatives | | | | / | / | / |

| | | | | | | | | |
|---|---|---|---|---|---|---|---|---|
| | doi: 10.1016/j.drugalcdep.2013.03.001. Epub 2013 Apr 3. PMID: 23561823 | Opioids | 3: high | | 2):352-61.001. Epub 2013 Apr 3. PMID: 23561823 | / | / | / |
| Life events<br><br>**The Social Readjustment Rating Scale**<br><br>43 items (Holmes et al. 1967)<br>44 items (Wallace et al. 2023) | Holmes, T.H., Rahe, R.H., 1967. The social readjustment rating scale. J. Psychosomatic Res. 11, 213e218 | / | 0–1871<br>-<br><150: low risk<br>150– 300: medium risk<br>>300: high risk | A higher score indicates that the individual has been confronted with more and/or worse stressors, which may expose them to a greater risk of deteriorating health. | Wallace D, Cooper NR, Sel A, Russo R. The social readjustment rating scale: Updated and modernised. PLoS One. 2023 Dec 18;18(12):e0295943. doi: 10.1371/journal.pone.0295943. PMID: 38109368; PMCID: PMC10727443. | / | David K. Harmon, Minoru Masuda, Thomas H. Holmes, The social readjustment rating scale: A cross-cultural study of Western Europeans and Americans, Journal of Psychosomatic Research, Volume 14, Issue 4, 1970, Pages 391-400, ISSN 0022-3999 | / |
| Personality traits<br><br>**Big Five Inventory**<br><br>44 items (John & Srivastava 1999)<br>45 items (Plaisant et al. 2010) | John, O. P., & Srivastava, S. (1999). The Big-Five trait taxonomy: History, measurement, and theoretical perspectives. In L. A. Pervin & O. P. John (Eds.), Handbook of personality: Theory and research (Vol. 2, pp. 102–138). New York: Guilford Press | Agreeableness | 1–5 | A higher score indicates a higher expression of the trait. | Husain, W., Haddad, A.J., Husain, M.A. *et al.* Reliability generalization meta-analysis of the internal consistency of the Big Five Inventory (BFI) by comparing BFI (44 items) and BFI-2 (60 items) versions controlling for age, sex, language factors. *BMC Psychol* 13, 20 (2025). https://doi.org/10.1186/s40359-024-02271-x | 0.73 | Plaisant O, Courtois R, Réveillère C, Mendelsohn G, John OP. Validation par analyse factorielle du Big Five Inventory français (BFI-Fr). Analyse convergente avec le NEO-PI-R. Annales médico-psychologique, 168, 97-106, 2010 | 0.75 |
| | | Openness | 1–5 | | | 0.77 | | 0.74 |
| | | Consciousness | 1–5 | | | 0.80 | | 0.80 |
| | | Extraversion | 1–5 | | | 0.80 | | 0.82 |
| | | Neuroticism | 1–5 | | | 0.80 | | 0.82 |
| Stress level<br><br>**Perceived Stress Scale**<br><br>10 items | Cohen S, Kamarck T, Mermelstein R. A global measure of perceived stress. J Health Soc Behav. 1983 Dec;24(4):385-96. PMID: 6668417 | / | 0–40<br>-<br>0–13: low<br>14–26: moderate<br>27–40: high | A higher score indicates higher perceived stress. | Review of the Psychometric Evidence of the Perceived Stress Scale<br>Lee, Eun-Hyun<br>Asian Nursing Research, Volume 6, Issue 4, 121 - 127 | >.70 | Lesage FX, Berjot S, Deschamps F. Psychometric properties of the French versions of the Perceived Stress Scale. Int J Occup Med Environ Health. juin 2012;25(2):178-84 | 0.83 |
| Coping strategies<br><br>**Brief COPE Inventory**<br><br>28 items | Carver, C. S. (1997). You want to measure coping but your protocol's too long: Consider the Brief COPE. International Journal of | Social support | 1–4 | A higher score indicates a greater use of the coping strategy. | / | / | Baumstarck K, Alessandrini M, Hamidou Z, Auquier P, Leroy T, Boyer L. Assessment of coping: a new French four-factor structure of the brief COPE inventory. Health Qual | 0.82 |
| | | Problem solving | 1–4 | | / | / | | 0.74 |
| | | Avoidance | 1–4 | | / | / | | 0.64 |

| | | | | | | | | |
|---|---|---|---|---|---|---|---|---|
| | Behavioral Medicine, 4(1), 92–100 | Positive thinking | 1–4 | | / | / | Life Outcomes. 11 janv 2017;15:8 | 0.71 |
| Chronotype<br>**Composite Scale of Morningness**<br>13 items | Smith, C. S., Reilly, C., & Midkiff, K. (1989). Evaluation of three circadian rhythm questionnaires with suggestions for an improved measure of morningness. Journal of Applied Psychology, 74(5), 728–738 | / | 13–55<br>-<br><22: evening chronotype<br>22–44: intermediate chronotype<br>>44: morning chronotype | / | Smith, C. S., Reilly, C., & Midkiff, K. (1989). Evaluation of three circadian rhythm questionnaires with suggestions for an improved measure of morningness. Journal of Applied Psychology, 74(5), 728–738 | 0.87 | Caci H, Nadalet L, Staccini P, Myquel M, Boyer P. Psychometric properties of the French version of the composite scale of morningness in adults. Eur Psychiatry J Assoc Eur Psychiatr. sept 1999;14(5):284-90. | 0.85 |
| Sleepiness<br>**Epworth Sleepiness Scale**<br>8 items | Johns MW. A new method for measuring daytime sleepiness: The Epworth Sleepiness Scale. Sleep. 1 nov 1991;14(6):540-5. | / | 0–24<br>-<br>≥10: excessive daytime sleepiness | A higher score indicates higher daytime sleepiness. | Shahid A, Shen J, Shapiro CM. Measurements of sleepiness and fatigue. J Psychosom Res. 2010 Jul;69(1):81-9. doi: 10.1016/j.jpsychores.2010.04.001. PMID: 20630266 | 0.88 | Kaminska M, Jobin V, Mayer P, Amyot R, Perraton-Brillon M, Bellemare F. The Epworth Sleepiness Scale: Self-administration versus administration by the physician, and validation of a French version. Can Respir J. 2010 Mar-Apr;17(2):e27-34. doi: 10.1155/2010/438676. PMID: 20422065; PMCID: PMC2866221. | 0.88 |
| Sleep quality<br>**Pittsburgh Sleep Quality Index**<br>19 items | Buysse DJ, Reynolds CF 3rd, Monk TH, Berman SR, Kupfer DJ. The Pittsburgh Sleep Quality Index: A new instrument for psychiatric practice and research. Psychiatry Res. 1989 May;28(2):193-213. doi: 10.1016/0165-1781(89)90047-4. PMID: 2748771 | Total score | 0–21<br>-<br>≥5: sleep disturbances | A higher score indicates greater sleep disturbances. | Buysse DJ, Reynolds CF 3rd, Monk TH, Berman SR, Kupfer DJ. The Pittsburgh Sleep Quality Index: A new instrument for psychiatric practice and research. Psychiatry Res. 1989 May;28(2):193-213. doi: 10.1016/0165-1781(89)90047-4. PMID: 2748771 | 0.83 | Blais FC, Gendron L, Mimeault V, Morin CM. Evaluation de l'insomnie: validation de trois questionnaires [Evaluation of insomnia: validity of 3 questionnaires]. Encephale. 1997 Nov-Dec;23(6):447-53. French. PMID: 9488928. | 0.88 |
| | | Subjective sleep quality | 0–3 | A higher score indicates greater disturbances. | | / | | / |
| | | Sleep latency | 0–3 | | | / | | / |
| | | Sleep duration | 0–3 | | | / | | / |
| | | Habitual sleep efficiency | 0–3 | | | / | | / |
| | | Sleep disturbances | 0–3 | | | / | | / |
| | | Use of sleeping medication | 0–3 | | | / | | / |

| | | | | | | | | |
|---|---|---|---|---|---|---|---|---|
| | | Daytime dysfunction | 0–3 | | | / | | / |
| Fatigue<br><br>**Multidimensional Fatigue Inventory**<br><br>20 items | Smets E.M.A., Garssen B., Bonke B., Dehaes J.C.J.M., “The Multidimensional Fatigue Inventory (MFI) psychometric properties of an instrument to assess fatigue”, Journal of Psychosomatic Research, 1995;39:315-25 | General fatigue | 9–45 | A higher score indicates a higher level of fatigue | / | / | Gentile S, Delarozière JC, Favre F, Sambuc R, San Marco JL. Validation of the French « multidimensional fatigue inventory » (MFI 20). Eur J Cancer Care (Engl). mars 2003;12(1):58-64. | 0.92 |
| | | Mental fatigue | 6–30 | | / | / | | 0.84 |
| | | Reduced activity | 3–15 | | / | / | | 0.68 |
| | | Reduced motivation | 2–10 | | / | / | | 0.73 |
| Physical activity and sedentary behaviors<br><br>**ONAPS Physical Activity Questionnaire**<br>adapted version for students | Charles M, Thivel D, Verney J, Isacco L, Husu P, Vähä-Ypyä H, et al. Reliability and validity of the ONAPS Physical Activity Questionnaire in assessing physical activity and sedentary behavior in French adults. Int J Environ Res Public Health. 25 mai 2021;18(11):5643 | Vigorous activity | / | A higher score indicates a higher level of the corresponding dimension. | / | / | Charles M, Thivel D, Verney J, Isacco L, Husu P, Vähä-Ypyä H, et al. Reliability and validity of the ONAPS Physical Activity Questionnaire in assessing physical activity and sedentary behavior in French adults. Int J Environ Res Public Health. 25 mai 2021;18(11):5643. | / |
| | | Moderate activity | / | | / | / | | / |
| | | Light activity | / | | / | / | | / |
| | | Sedentary behavior | / | | / | / | | / |
| | | Active travel | / | | / | / | | / |
| Physical activity<br><br>**Rapid Assessment of Physical Activity Questionnaire**<br>adapted version for students | Topolski TD, LoGerfo J, Patrick DL, Williams B, Walwick J, Patrick MB. The rapid assessment of physical activity (RAPA) among older adults. Prev Chronic Dis. 2006 Oct;3(4):A118. Epub 2006 Sep 15. PMID: 16978493; PMCID: PMC1779282 | Vigorous activity | / | A higher score indicates a higher level of the corresponding dimension. | Topolski TD, LoGerfo J, Patrick DL, Williams B, Walwick J, Patrick MB. The rapid assessment of physical activity (RAPA) among older adults. Prev Chronic Dis. 2006 Oct;3(4):A118. Epub 2006 Sep 15. PMID: 16978493; PMCID: PMC1779282. | / | / | / |
| | | Moderate activity | / | | | / | / | / |
| | | Light activity | / | | | / | / | / |
| | | Strength activity | / | | | / | / | / |
| | | Flexibility activity | / | | | / | / | / |

#### Appendix 4. Physical fitness tests

The following set of physical tests will be performed under the supervision of a previously trained investigator and a physical activity instructor. Verbal encouragement will be provided to ensure optimal performance (1).

**Strength**

*Voluntary isometric maximal quadriceps force test.* Lower limb muscle torque will be assessed using an isometric quadriceps force test. The student will be seated with their hip and knee joints at 90° and their arms crossed over their chest. The right upper leg will be fixed to the chair to minimize extraneous movement, while the ankle will be attached to a dynamometer (DFS II, Chatillon Force Measurement, AMETEK STC, USA) (2,3). First, the student will perform a warm-up with a three-second intended extension at 10%, 30%, and 70% of their theoretical maximal voluntary isometric contraction. Second, two maximal isometric voluntary contractions (MIVC) will be performed, during which the student will be instructed to try to extend their leg as much as possible (2,3). The contractions will be separated by one minute of recovery to prevent muscle fatigue (4). The highest value will be used to determine the student's maximum isometric strength (in N) (5).

**Power**

*The squat jump test.* The student will adopt the starting squat position and hold it for three seconds before performing a vertical jump, with a 90° knee angle and their hands placed at their sides. The jump will be performed without a previous countermovement. Two or three warm-up jumps will be completed to ensure familiarization. After receiving the start signal, the student will perform two jumps, aiming to reach the maximum height, with a one-minute recovery period between each attempt. Jump heights will be recorded (in cm) using the Optojump system (Microgate Srl, Bolzano, Italy, (8)). The highest jump height (cm) will be used to estimate peak power, calculated using the Sayers equation: Peak Power (W) = 60.7 × jump height (cm) + 45.3 × body mass (kg) – 2055. Relative peak power will then be obtained by dividing peak power (W) by body mass (kg).

**Flexibility**

*Shoulder stretch test*. The shoulder stretch test is a functional test that will be used to assess overall shoulder mobility, particularly in the glenohumeral joints, as well as the symmetry and muscular balance of the scapulothoracic complex. The participant will place one hand behind their head (external rotation and abduction) and the other behind their back (internal rotation and adduction), with the aim of bringing the fingers closer together or touching. This test is included in the Functional Movement Screen (FMS) battery and can identify limitations in joint range of motion, bilateral imbalances, or functional restrictions that may increase the risk of injury. Measurements will be based on the distance between the hands and may be further categorized on a scale from 5 to 1, as follows: 5 - overlapping their fingers , 4 - fingers touching, 3 - fingers barely touching, 2 - fingers not touching, 1 - and not touching their own shoulder (1). Its inter- and intra-rater reliability are high, with intraclass correlation coefficients generally above 0.90 (11,12). It is a simple, rapid, and relevant tool for clinical screening (13).

*Sit-and-reach test*. The sit-and-reach test is a standardized test used to assess hamstring flexibility and, to a lesser extent, lumbar mobility (9). The participant will sit with their knees fully extended and their bare feet flat and dorsiflexed against a box or flat surface. The participant will align their fingers with one hand resting on top of the other while they perform a slow, active forward flexion of the trunk. This movement progressively stretches the gastrocnemius, hamstrings, hip external rotators, and erector spinae muscles. The participant will flex in their full range of motion by a controlled concentric contraction of the hip flexors and abdominal muscles, reaching the maximum limit without pain. At the end of the movement, an isometric contraction will be held for approximately two seconds before the distance between the fingers and toes is measured. The measurement was recorded as a positive value when the participant was able to reach beyond the toes (corresponding to the distance between the fingertips and the toes), and as a negative value when the participant was unable to reach the toes.This score reflects overall posterior chain flexibility, with particular emphasis on hamstring extensibility (10).

**Endurance**

*Cardiorespiratory fitness test*. The 20-m shuttle run test is a common indirect method for estimating individual cardiorespiratory fitness (14). In this test, participants run back and forth between two lines 20 metres apart, gradually increasing their speed. The highest speed achieved during the test is used to predict the maximal oxygen uptake ($VO_2max$) (9,14–16). Due to local constraints the modified 10-m version will be used in the current study (16,18). Specifically, the lines will be set 10 metres apart, and the starting speed will be 6.5 km/h. The running pace will increase at regular intervals (+0.5km/h per minute), guided by an audio signal (Pack Sportbeeper Pro, CE, France). The test will stop when the student can no longer maintain the required speed, indicating their Maximal Aerobic Speed (MAS).

Each student's heart rate will be assessed during and after the shuttle test using a Polar A200 device connected to a Polar H7 chest strap sensor. Their heart rate will be recorded immediately after they stop, as well as one and three minutes after they stop. Heart rate recovery (HRR) will be measured in beats per minute by subtracting the heart rate at one (HRR1) and three minutes (HRR3) of recovery from the heart rate at the time of stopping (HRS). The estimated $VO_2max$ (mL/kg/min) will be calculated using an adjusted equation for this distance ($VO_{2max}$ = 8.0574 * 10m-SRT speed - 32.64).

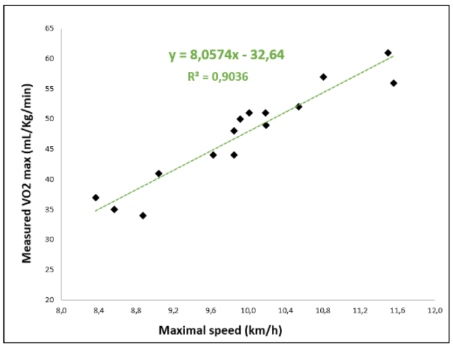

#### Appendix 5. Module-related activities

While waiting for individual meetings with the experts, students will be permitted to engage in module-related activities. These activities will be supervised by an experimenter to ensure students' well-being. Interactions during sessions will be promoted by encouraging students to actively participate in the activities outlined below.

**Session 1:** The RESPIRELAX+ interface will be explained. The experimenter will then explain to the student how to fill out the stress diary, and for practice, the student will fill in the diary entry for that day. They will be instructed to continue updating it daily until the next session. The experimenter will provide information about the local student health service. Finally, students will be given a certificate of attendance.

**Session 2:** The students and the experimenter will discuss the stress diary completed during the previous session and review any difficulties they encountered while filling it out.

**Session 3:** The experimenter will recommend that students install a blue light filter on their phones. The experimenter will then provide the students with the sleep diary. They will fill it out for the previous night. Students will be given sleep masks and earplugs. Finally, students will watch a video titled "To Sleep, Perchance to Dream: Crash Course Psychology #9" (https://www.youtube.com/watch?v=rMHus-0wFSo&ab_channel=CrashCourse).

**Session 4:** Medical students will receive advice on managing night shifts, while doctoral students will discuss parallels with social jet lag during conferences or vacations. The experimenter and students will review the previous sleep diaries in order to address any difficulties students might have had in completing them. Each student will be asked to undergo a bioimpedance analysis (Biody XpertZM 3; Aminogram SAS, La Ciotat, France). The physical activity diary will then be explained, and students will complete it for the previous day. They will also be invited to download Apple Health or Google Fit to track their daily steps. Finally, students will watch the video "Muscles, Part 1 – Muscle Cells: Crash Course Anatomy & Physiology #21" (https://youtu.be/Ktv-CaOt6UQ?si=z2IO5eJRyI33J8_U).

**Session 5:** The physical activity diaries completed during the previous session will be reviewed as described above. A new bioimpedance analysis and a handgrip test will be proposed. The handgrip test assess forearm strength using a Jamar® Smart Hand Dynamometer (MD; Performance Health Supply, Cedarburg, WI 53012). The test will be conducted in a standardized manner: students will be seated with their feet flat on the floor, shoulders adducted, elbows flexed at 90°, forearms in a neutral position, and wrists slightly extended. The handle will be adjusted for optimal comfort. Students will be asked to squeeze as hard as possible for three to five seconds without compensating with their trunk or shoulder. Two to three trials will be performed for each hand, and students will take 1-min breaks between attempts. The highest value will be recorded as the maximal isometric strength (in N). Finally, students will watch the video "Muscles, Part 2 – Organismal Level: Crash Course Anatomy & Physiology #22" (https://youtu.be/I80Xx7pA9hQ?si=1VQ8baxjyDJHaMcF). *Handgrip test.*

**Session 6:** Students will watch the video "Metabolism & Nutrition, Part 1: Crash Course Anatomy & Physiology #36" (https://youtu.be/fR3NxCR9z2U?si=YBC6XyfPjAYK73E0) and "Metabolism & Nutrition, Part 2: Crash Course Anatomy & Physiology #37" (https://youtu.be/kb146Y1igTQ?si=QD98PUKMV-PlgsnT).

#### Appendix 6. Set of articles

To become a PROMESS-Group expert, individuals who will conduct the session should have read the following articles regarding the stress, sleep and physical activity module.

| Stress | Sleep | Physical activity |
|---|---|---|
| Dyrbye et al. (2005)<br>**Medical student distress: causes, consequences, and proposed solutions**<br>Mayo Clin Proc.<br>https://doi.org/10.4065/80.12.1613 | Baranwal et al. (2023)<br>**Sleep physiology, pathophysiology, and sleep hygiene**<br>Prog Cardiovasc Dis.<br>https://doi.org/10.1016/j.pcad.2023.02.005 | Gao et al. (2024)<br>**Occupational sitting time, leisure physical activity, and all-cause and cardiovascular disease mortality**<br>JAMA Network Open<br>https://doi.org/10.1001/jamanetworkopen.2023.50680 |
| LeBlanc. (2009)<br>**The Effects of acute stress on performance: Implications for health professions education**<br>Acad Med.<br>https://doi.org/10.1097/acm.0b013e3181b37b8f | Ohayon et al. (2017)<br>**National Sleep Foundation's sleep quality recommendations: first report**<br>Sleep Health.<br>https://doi.org/10.1016/j.sleh.2016.11.006 | Bull et al. (2020)<br>**World Health Organization 2020 guidelines on physical activity and sedentary behaviour**<br>Br J Sports Med.<br>https://doi.org/10.1136/bjsports-2020-102955 |
| Shapiro et al. (2000)<br>**Stress management in medical education: a review of the literature**<br>Acad Med.<br>https://doi.org/10.1097/00001888-200007000-00023 | Windred et al. (2023)<br>**Sleep regularity is a stronger predictor of mortality risk than sleep duration: A prospective cohort study**<br>Sleep<br>https://doi.org/10.1093/sleep/zsad253 | Tremblay et al. (2017)<br>**Sedentary Behavior Research Network (SBRN) – Terminology Consensus Project process and outcome**<br>**International Journal of Behavioral Nutrition and Physical Activity**<br>International Journal of Behavioral Nutrition and Physical Activity<br>https://doi.org/10.1186/s12966-017-0525-8 |
| Shiralkar et al. (2013)<br>**A Systematic Review of Stress-Management Programs for Medical Students**<br>Acad Psychiatry<br>https://doi.org/10.1176/appi.ap.12010003 | Azad et al. (2015)<br>**Sleep disturbances among medical students: a global perspective**<br>J Clin Sleep Med.<br>https://doi.org/10.5664/jcsm.4370 | Peleias et al. (2017)<br>**Leisure time physical activity and quality of life in medical students: results from a multicentre study**<br>BMJ Open Sport Exerc Med.<br>https://doi.org/10.1136/bmjsem-2016-000213 |

| | | |
|---|---|---|
| Fincham et al. (2023)<br>**Effect of breathwork on stress and mental health: A meta-analysis of randomised-controlled trials**<br>Sci Rep.<br>https://doi.org/10.1038/s41598-022-27247-y | Seoane et al. (2020)<br>**Sleep disruption in medicine students and its relationship with impaired academic performance: A systematic review and meta-analysis**<br>Sleep Med Rev.<br>https://doi.org/10.1016/j.smrv.2020.101333 | Hoffmann et al. (2025)<br>**Prevalence and correlates of self-reported and accelerometer-determined sedentary behavior and physical activity of German university students: cross-sectional results of the SmartMoving study**<br>BMC Public Health<br>https://doi.org/10.1186/s12889-025-24378-5 |
| Schlatter et al. (2022)<br>**Effects of relaxing breathing paired with cardiac biofeedback on performance and relaxation during critical simulated situations: a prospective randomized controlled trial**<br>BMC Med Educ.<br>https://doi.org/10.1186/s12909-022-03420-9 | Perotta et al. (2021)<br>**Sleepiness, sleep deprivation, quality of life, mental symptoms and perception of academic environment in medical students**<br>BMC Med Educ.<br>https://doi.org/10.1186/s12909-021-02544-8 | Al-Drees et al. (2016)<br>**Physical activity and academic achievement among the medical students: A cross-sectional study**<br>Medical Teacher<br>http://dx.doi.org/10.3109/0142159X.2016.1142516 |
| Schlatter et al. (2022)<br>**Personality traits affect anticipatory stress vulnerability and coping efficacy in occupational critical care situations**<br>Sci Rep - https://doi.org/10.1038/s41598-022-24905-z | Bani Issa et al. (2023)<br>**Evaluation of the effectiveness of sleep hygiene education and FITBIT devices on quality of sleep and psychological worry: a pilot quasi-experimental study among first-year college students**<br>Front Public Health.<br>https://doi.org/10.3389/fpubh.2023.1182758 | Leuchter et al. (2022)<br>**Relationship between exercise intensity and stress levels among U.S. medical students**<br>Medical Education Online<br>https://doi.org/10.1080/10872981.2022.2027651 |
| Le Saux et al. (2024)<br>**Association of personality traits with the efficacy of stress management interventions for medical students taking objective structured clinical examinations**<br>Acad Med.<br>https://doi.org/10.1097/ACM.0000000000005714 | Chandler et al. (2022)<br>**Improving university students' mental health using multi-component and single-component sleep interventions: A systematic review and meta-analysis**<br>Sleep Med.<br>https://doi.org/10.1016/j.sleep.2022.09.003 | Plotnikoff et al. (2015)<br>**Effectiveness of interventions targeting physical activity, nutrition and healthy weight for university and college students: A systematic review and meta-analysis**<br>Int J Behav Nutr Phys Act<br>https://doi.org/10.1186/s12966-015-0203-7 |

| | | |
|---|---|---|
| Barret et al. (2024)<br>**Associations of coping and health-related behaviors with medical students' well-being and performance during objective structured clinical examination**<br>Sci Rep.<br>https://doi.org/10.1038/s41598-024-61800-1 | Shriane et al. (2023)<br>**Healthy sleep practices for shift workers: consensus sleep hygiene guidelines using a Delphi methodology**<br>Sleep.<br>https://doi.org/10.1093/sleep/zsad182 | Thorndike et al. (2014)<br>**Activity monitor intervention to promote physical activity of physicians-in-training: Randomized controlled trial**<br>PLoS One<br>https://doi.org/10.1371/journal.pone.0100251 |
| Métais et al. (2025)<br>**Determining the influence of an intervention of stress management on medical students' levels of psychophysiological stress: the protocol of the PROMESS-Stress clinical trial**<br>BMC Medical Education<br>https://doi.org/10.1186/s12909-024-06344-8 | Ruet et al. (2025)<br>**Determining the influence of a sleep improvement intervention on medical students' sleep and fatigue: protocol of the PROMESS-Sleep clinical trial**<br>BMC Medical Education<br>https://doi.org/10.1186/s12909-024-06422-x | Schlatter et al. (2025)<br>**Reducing Sedentary Behavior and Promoting Physical Activity in Medical Students: Protocol of the PROMESS-Physical Activity Clinical Trial**<br>https://doi.org/10.21203/rs.3.rs-7441836/v1 |

#### Appendix 7. Visual analogue scales

Students will answer several 100-mm visual analogue scales (VASs). Each VAS will prompt the student to move a cursor to indicate their response. The cursor will start at the 0 position for each question.

| Stress - Session 1 - 2 - 3 | Sleep - Session 3 - 4 - 5 | Physical activity - Session 4 - 5 - 6 |
|---|---|---|
| **VAS - stressors quantity**<br>"In the past two weeks, how many stressful situations have you encountered?"<br>Ranging from 0 (maximum/extreme) to 100 (zero). | **VAS - sleep quantity**<br>"In the past 2 weeks, how would you characterize your sleep quantity?"<br>Ranging from 0 (largely insufficient) to 100 (largely sufficient) | **VAS - physical fitness**<br>"Over the past two weeks, how would you characterize your level of physical fitness?"<br>Ranging from 0 (bad shape) to 100 (excellent shape). |
| **VAS - stress quantity**<br>"In the past two weeks, how would you characterize your stress level?"<br>Ranging from 0 (maximum stress) to 100 (no stress at all). | **VAS - sleep quality**<br>"In the past 2 weeks, how would you characterize your sleep quality?"<br>Ranging from 0 (the worst quality) to 100 (the best quality). | **VAS - physical activity**<br>"Over the past two weeks, how would you characterize your level of physical activity?"<br>Ranging from 0 (completely inactive) to 100 (extremely active). |
| **VAS - stress quality**<br>"Over the past two weeks, how would you characterize your stress?"<br>Ranging from 0 (very negative feeling) to 100 (very positive feeling). | **VAS - fatigue**<br>"Over the past 2 weeks, how would you characterize your physical and mental fatigue level?"<br>Ranging from 0 (extreme fatigue) to 100 (no fatigue at all). | **VAS - sedentary behavior**<br>"Over the past two weeks, how would you characterize your level of sedentary behavior?"<br>Ranging from 0 (extremely sedentary behavior) to 100 (no sedentary behavior). |
| **VAS - stress coping**<br>"Over the past two weeks, how have you managed stressful situations?"<br>Ranging from 0 (very bad stress coping) to 100 (excellent stress coping). | | |

Appendix 8. Stress, sleep, and physical activity diaries

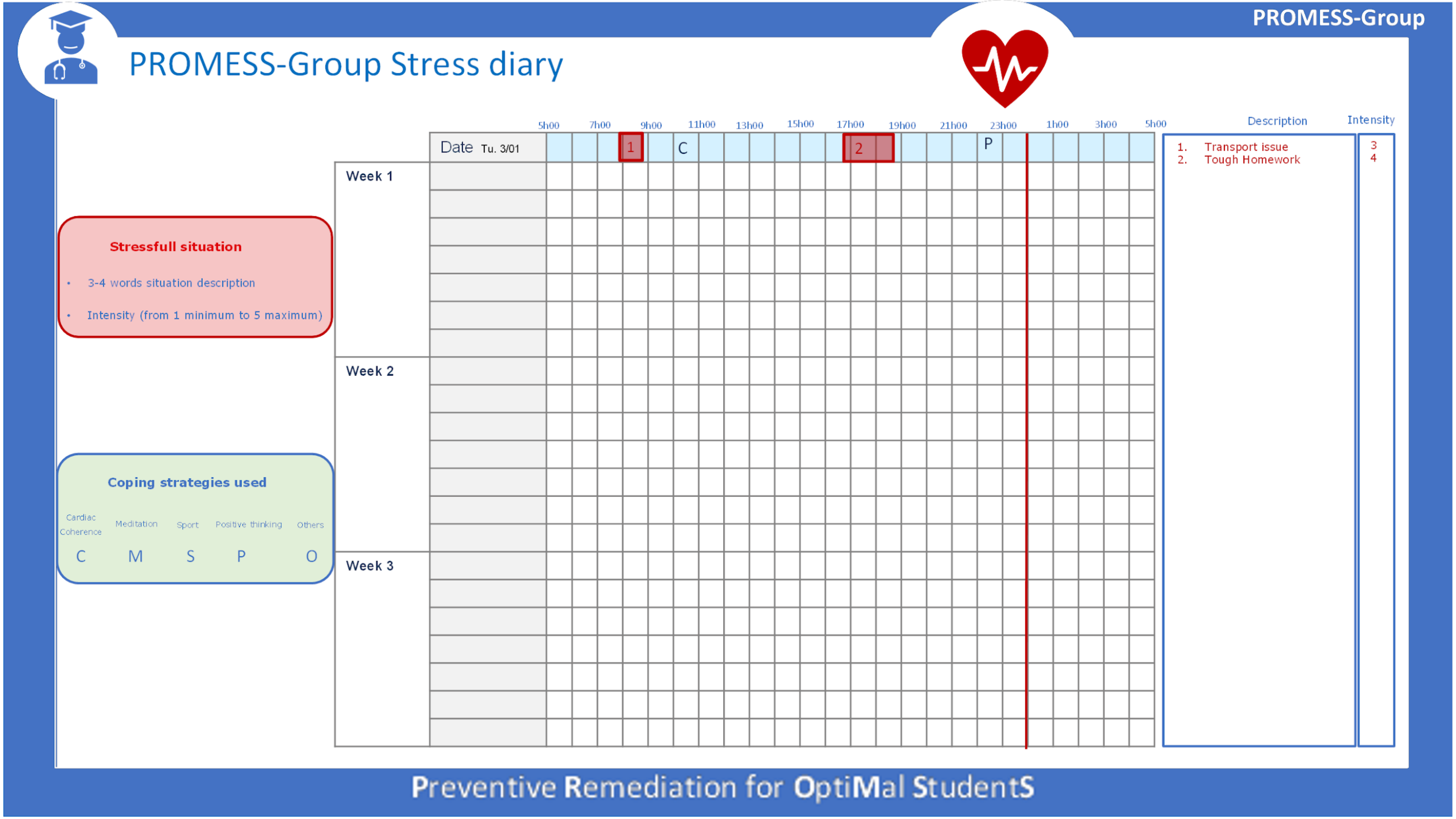

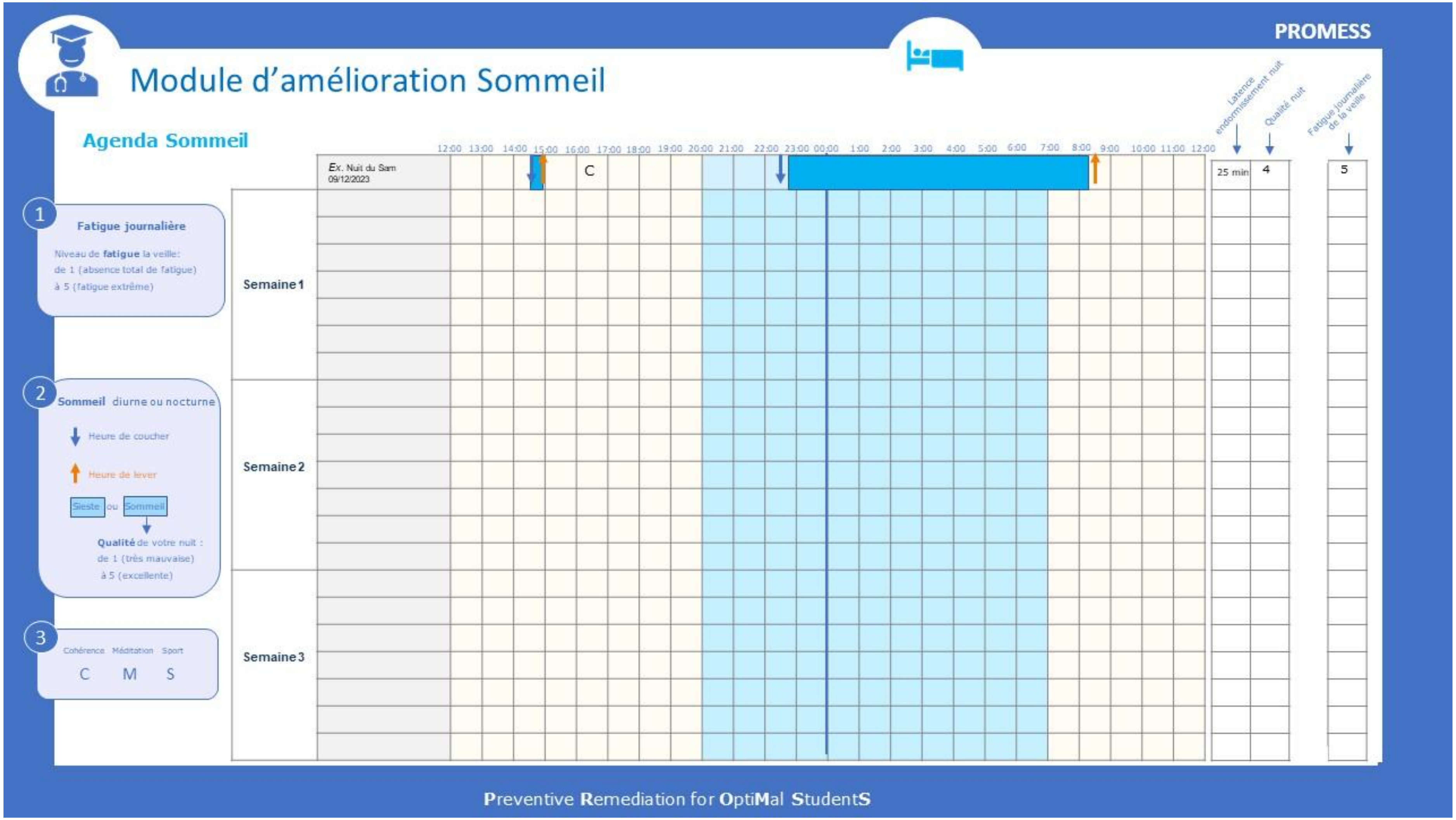
PROMESS
Module d'amélioration Sommeil
Agenda Sommeil
Latence endormissement nuit
Qualité nuit
Fatigue journalière de la veille
12:00 13:00 14:00 15:00 16:00 17:00 18:00 19:00 20:00 21:00 22:00 23:00 00:00 1:00 2:00 3:00 4:00 5:00 6:00 7:00 8:00 9:00 10:00 11:00 12:00
Ex. Nuit du Sam 09/12/2023
C
25 min
4
5
1
Fatigue journalière
Niveau de fatigue la veille:
de 1 (absence total de fatigue)
à 5 (fatigue extrême)
2
Sommeil diurne ou nocturne
Heure de coucher
Heure de lever
Sieste ou Sommeil
Qualité de votre nuit :
de 1 (très mauvaise)
à 5 (excellente)
3
Cohérence Méditation Sport
C M S
Semaine 1
Semaine 2
Semaine 3
Preventive Remediation for OptiMal StudentS

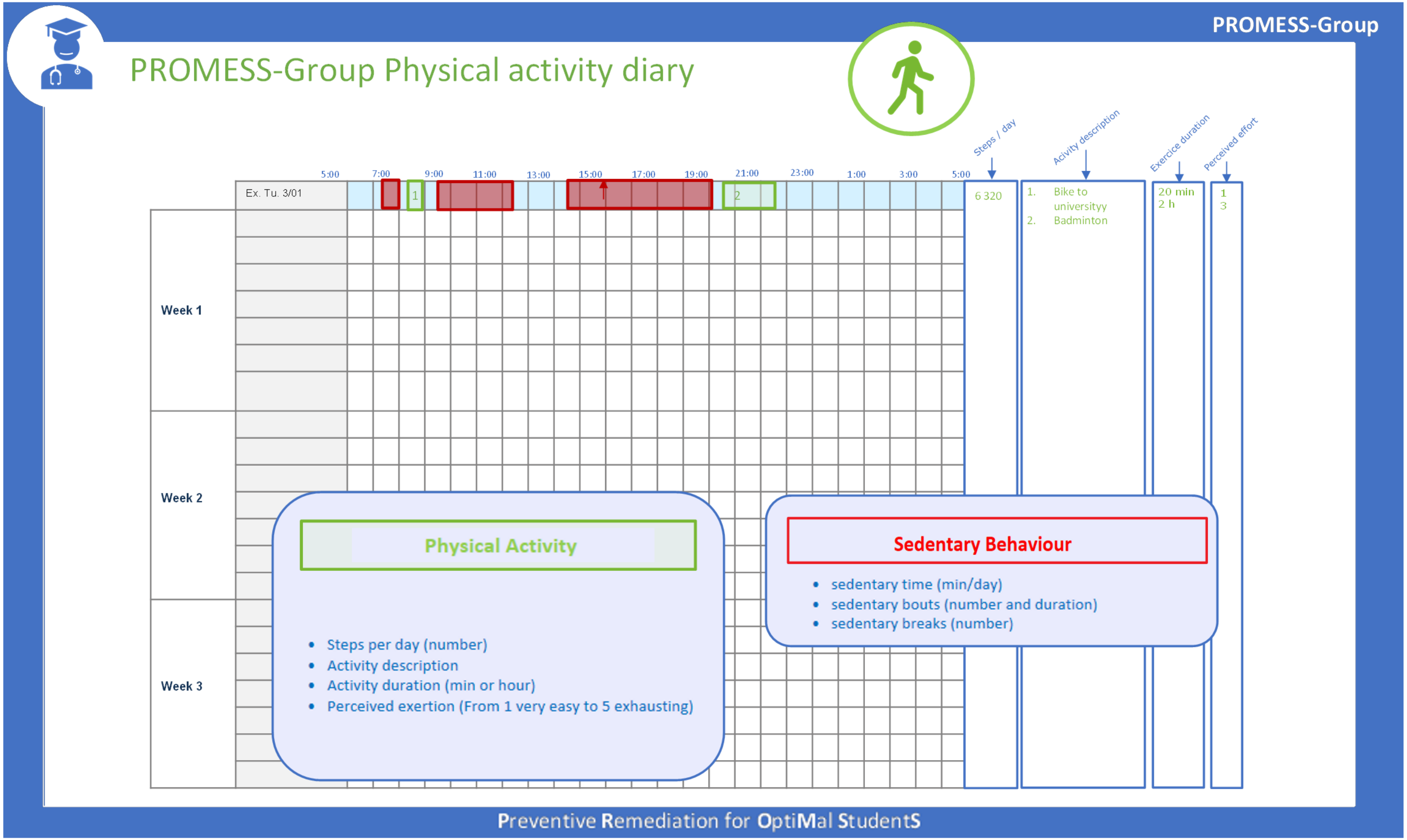
PROMESS-Group
PROMESS-Group Physical activity diary
Steps / day
Acivity description
Exercice duration
Perceived effort
5:00
7:00
9:00
11:00
13:00
15:00
17:00
19:00
21:00
23:00
1:00
3:00
5:00
Ex. Tu. 3/01
1
2
6 320
1. Bike to universityy
2. Badminton
20 min
2 h
1
3
Week 1
Week 2
Week 3
Physical Activity
Steps per day (number)
Activity description
Activity duration (min or hour)
Perceived exertion (From 1 very easy to 5 exhausting)
Sedentary Behaviour
sedentary time (min/day)
sedentary bouts (number and duration)
sedentary breaks (number)
Preventive Remediation for OptiMal StudentS

#### Appendix 9. Lists of advice

During sessions, the PROMESS-Group experts will provide students with specific advice and goals based on the following predefined lists regarding stress, management, sleep and physical activity. These lists are based on well-established knowledge and interventions in their respective fields; additional references are available (19–21). To ensure the list fit the specific needs of students, the list was validated by a sample of students and university staff members working in the health department during a prior workshop (22). The experts will use a 5-point Likert scale to report whether each piece of advice 1) was not mentioned, 2) was mentioned but not directly advised, 3) was mentioned and recommended, 4) was set as a goal, and 5) was mentioned with positive reinforcement performed.

| **Stress** | **Sleep** | **Physical activity** |
|---|---|---|
| Fill in the stress diary. | Fill in the sleep diary. | Fill in the physical activity diary. |
| Daily habits<br>- Take breaks<br>- Practice positive thinking<br><br>Relaxation methods<br>- Practice slowed breathing/cardiac coherence<br>- Practice meditation/mindfulness | Daytime habits<br>- Anti-blue light filter on screens<br>- Restricted screen use<br>- Restricted alcoholic consumption<br>- No energy drink or coffee intake after 4:00 p.m.<br>- Regular meal intake<br>- Eat or have a break outside to maximize sunlight exposure | Daily habits<br>- Check and/or increase the number of daily steps<br>- Take the stairs more often<br>- Increase walking and/or cycling |
| Social support<br>- Reinforce positive social relationships<br>- Participate in students' associative activities<br>- Participate in university health service activities<br><br>Physical activity<br>- Perform physical activity<br>- Participate in structured physical activities offered by the university | Nighttime habits<br>- Sleep for seven to nine hours nightly<br>- Regularize bedtime<br>- Total obscurity (have a sleep mask on)<br>- Silence (earplugs, devices switched off)<br>- No phone next to the bed<br>- No work inside the bed<br>- Go to sleep when tired<br>- Read before going to bed<br>- Having a bedtime routine<br>- Control the bedroom's temperature<br>- Relaxation technique (e.g., meditation and mindfulness, slow-paced breathing, cardiac coherence, guided imagery, muscle relaxation) | Physical activity<br>- Perform one moderate physical activity session per week<br>- Perform one intense physical activity session per week<br>- Gradually increase the duration of your sports activity sessions |
| Other<br>- List and reflect on coping behaviors<br>- Engage in artistic or manual activities<br>- Spend time in nature | Daytime sleep<br>- Power nap (10 to 20 minutes)<br>- Long nap (more than 30 minutes) | Sedentary behaviors<br>- Break up sedentary lifestyle free time (screen time, sitting time, etc.)<br>- Break up a sedentary lifestyle by moving around while sitting (metro, bus, etc.) |

#### Appendix 10. Pedagogical content examples

1) Pedagogical content from stress sessions

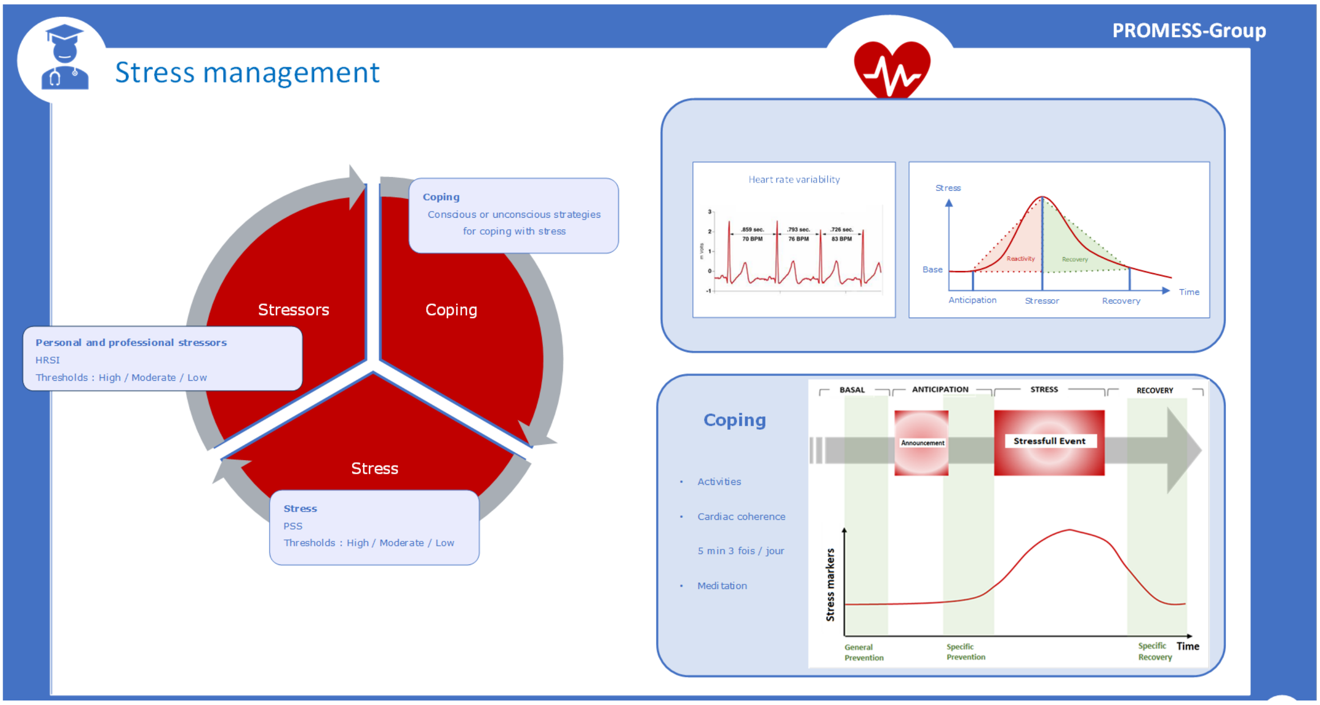


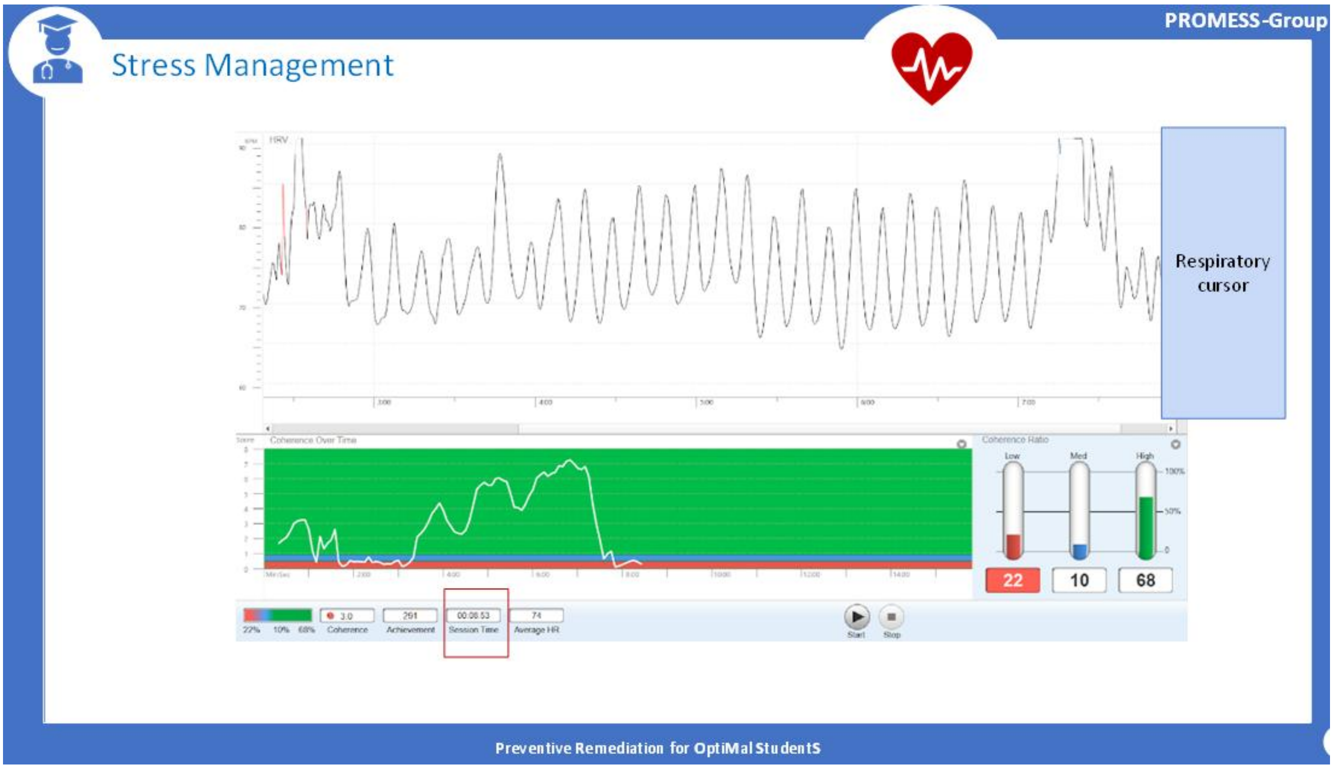

PROMESS-Group

### Stress Management

##### The Big Five Personality Traits

O (Openness to Experience): openness, originality, intellectual curiosity

C (Conscientiousness): conscientiousness, constraint, self-control

E (Extraversion): extraversion, energy, enthusiasm

A (Agreeableness): agreeableness, altruism, affection

N (Neuroticism): neuroticism, negative affectivity, nervousness

##### Relationships between Personality Traits and Stress Sensitivity

| | Traits | Sensibilité au stress |
|---|---|---|
| Openness to Experience | O+ | Limited literature |
| Conscientiousness | C+ | Good adherence to coping strategies (protective behaviors)<br>Caution: when associated with high Neuroticism → increased stress vulnerability |
| Extraversion | E+ | Stress résilience |
| Agreeableness | A+ | Limited literature |
| Neuroticism | N+ | Stress vulnerability |

Preventive Remediation for OptiMal StudentS

PROMESS-Group

### Stress management

**Coping**

ACTIVE RESOLUTION
Planning
Active efforts to reduce stress

POSITIVE THINKING
Humor
Acceptance
Positive reframing

AVOIDANCE
Denial
Self-criticism
Substance use
Behavioral disengagement
Distraction

SOCIAL SUPPORT
Seeking help/advice
Emotional support
Complaints
Religion

##### Relationships between Coping and Stress Sensitivity

| | Scores | Sensibility to stress |
|---|---|---|
| Active resolution | AR+ | Stress resilience |
| Positive Thinking | PT+ | Stress resilience |
| Avoidance | A+ | Stress vulnerability |
| Social support | SS+ | Depends on context |

Preventive Remediation for OptiMal StudentS

2) Pedagogical content from sleep sessions

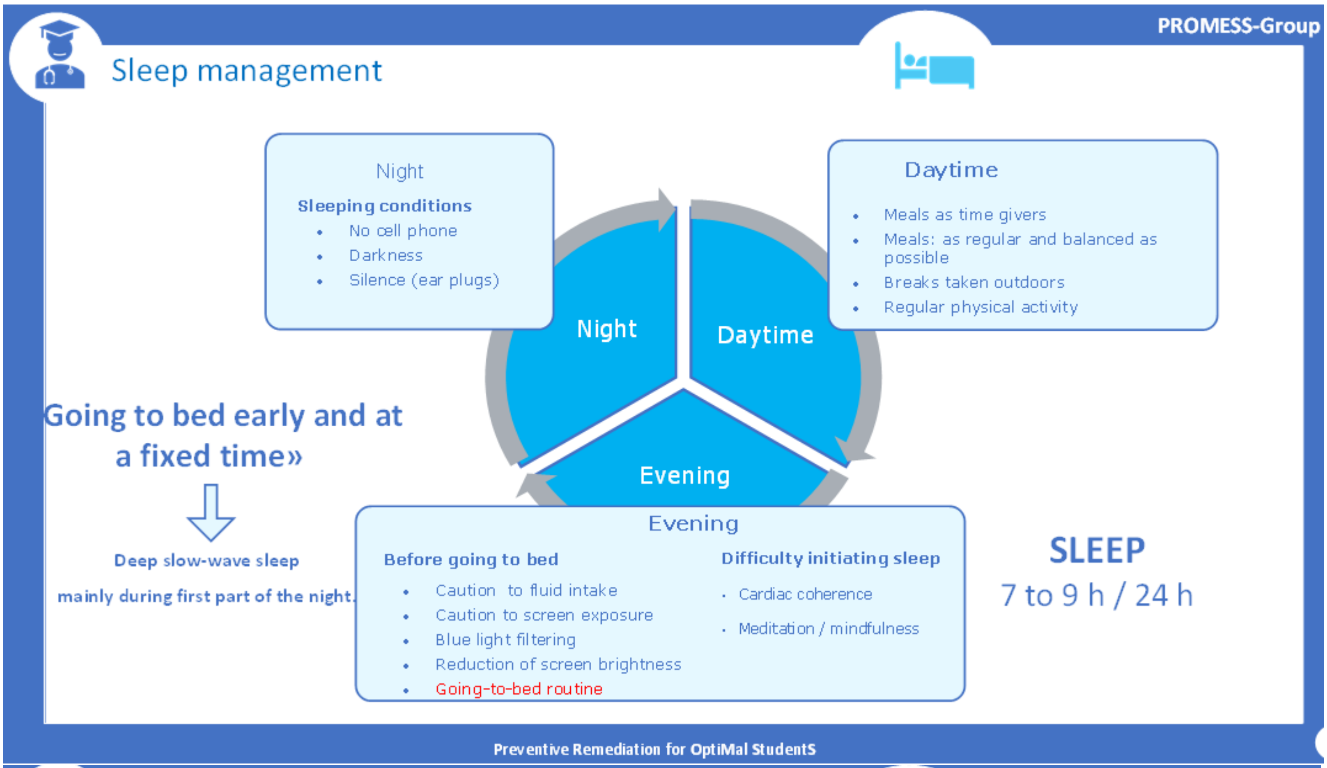


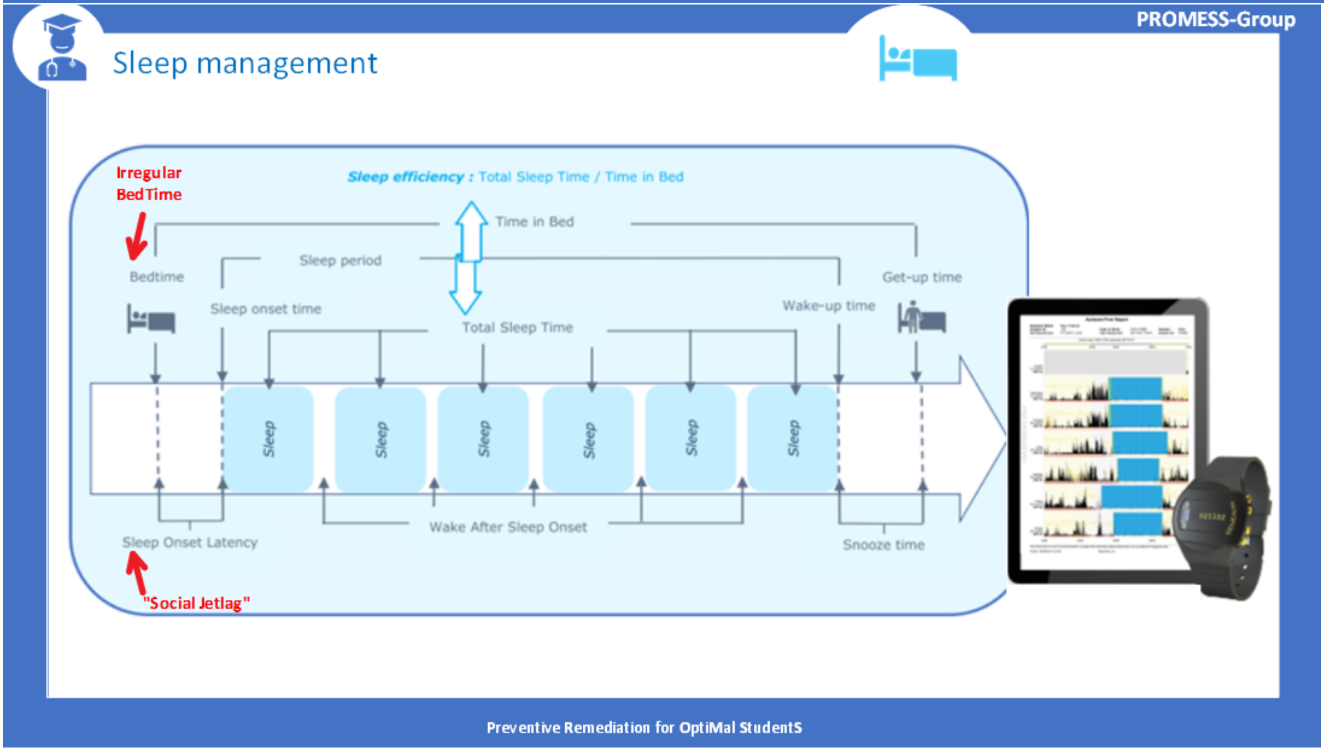

PROMESS-Group

#### Sleep Management

**Actigraphy :**

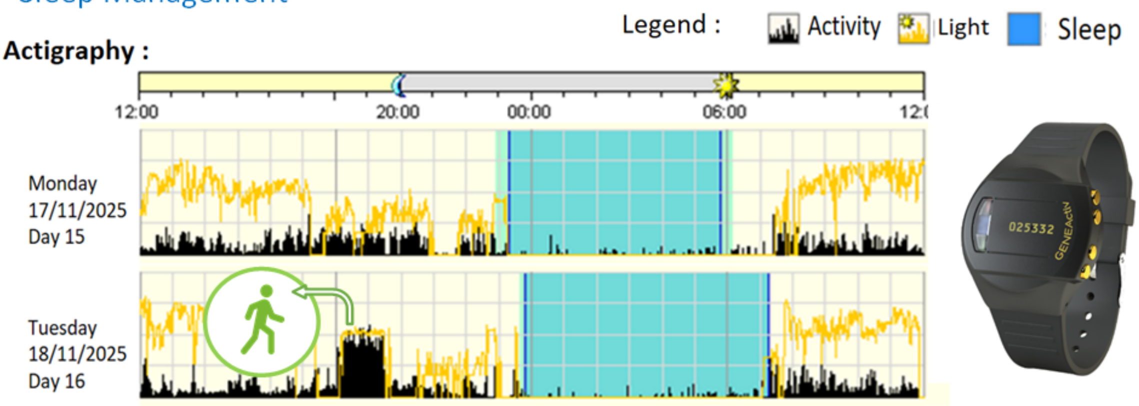


**Personnal statistics :**

| | BedTime | Wake-up Time | Time in bed (hours) | Total Sleep Time (hours) | Sleep onset Time (minutes) | Sleep efficiency (Percentage) | Wake after Onset (minutes) | Awakenings |
|---|---|---|---|---|---|---|---|---|
| Min | 16:40:00 | 01:35:00 | 6:20:00 | 3:15:00 | 0,00 | 47,45 | 0,00 | 0 |
| Max | 06:17:00 | 10:03:00 | 17:13:00 | 17:11:00 | 71,00 | 99,81 | 0,00 | 0 |
| Moy | 23:02:27 | 07:28:15 | 8:39:12 | 8:02:21 | 14,59 | 92,63 | 0,00 | 0,00 |
| | | | | Insufficient : > 7h | Excessive : > 30 min<br>Pathologic: < 5 min | Insufficient : < 85 %<br>Alarming : < 80 % | Excessive : > 40 min | |

Preventive Remediation for OptiMal StudentS

3) Pedagogical content from physical activity sessions

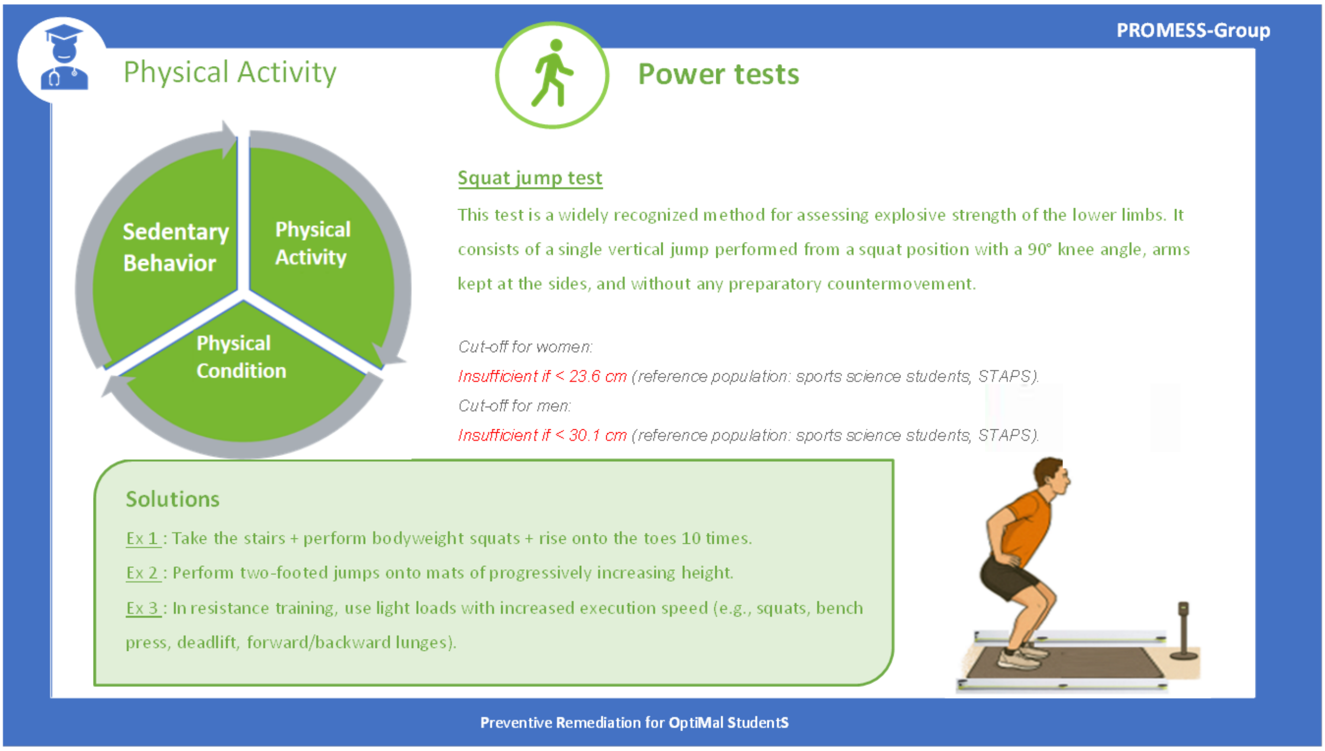


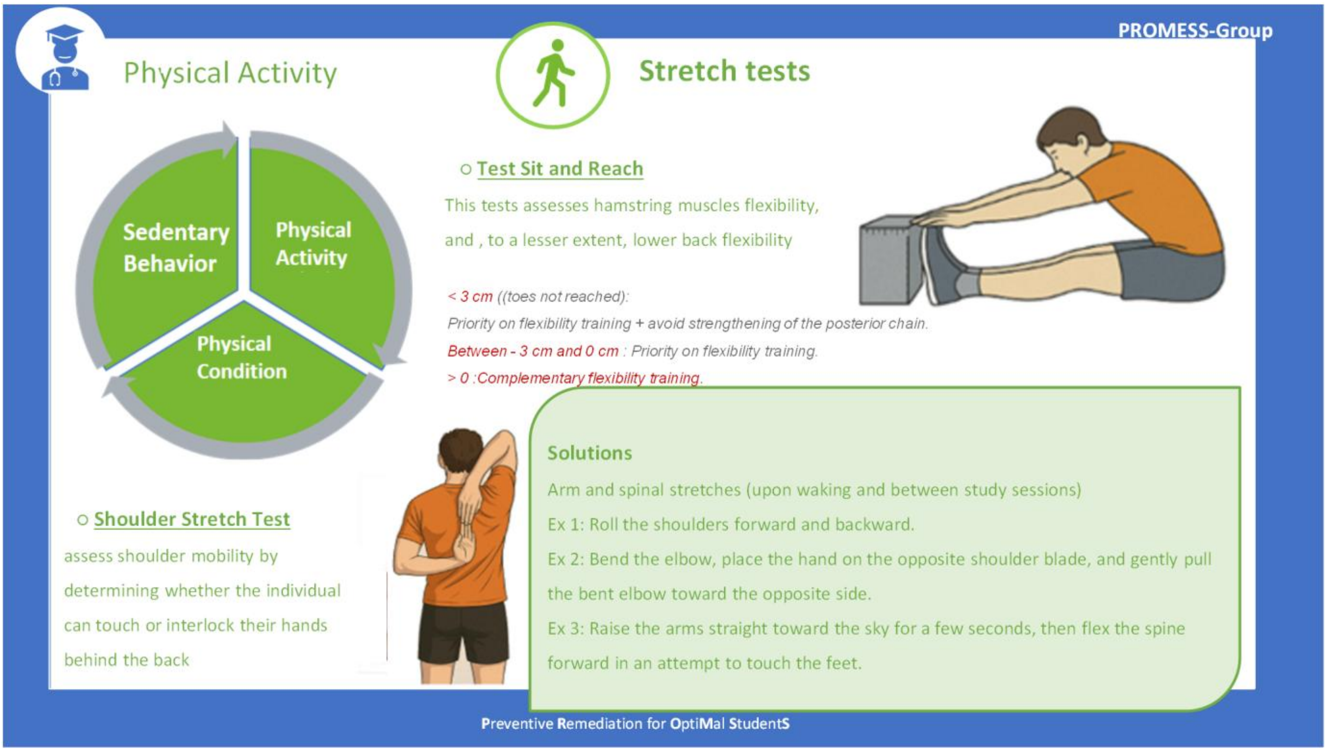

PROMESS-Group

#### Physical Activity

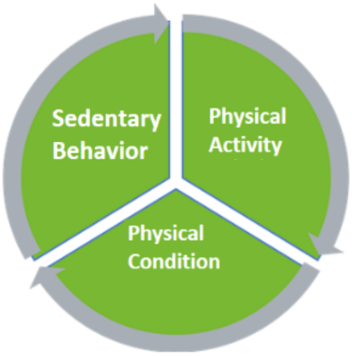


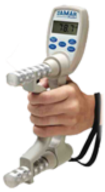

#### Strength

- **Voluntary isometric maximal quadriceps force test**

Assessed using an isometric quadriceps strength test with a dynamometer. Comparison method based on a threshold adjusted for age, sex, and body weight.:

*Women cut-ooff: ( 66,37 - (0,87 * age(in years) +(49,7 * 0) + (0,96 * weight(in kg))) / 0,4*

*Men cut-off: ( 66,37 - (0,87 * age(in years)) + (49,7 * 1) + (0,96 * weight(in kg))) / 0,4*

→ *Insufficient if force (N) is below the threshold*

##### Solutions

Ex 1 : Perform muscle-strengthening exercises: wall sit, core planks, abdominal exercises.

Ex 2 : Perform 10 squats during an active break. Mountain climber exercise.

Ex 3 : In resistance training: heavy loads for 10 repetitions (e.g., squats, bench press, deadlift, forward/backward lunges).

- **Handgrip test**

The handgrip test assesses the static muscular strength of the forearms and hands.

Preventive Remediation for OptiMal StudentS

PROMESS-Group

#### Physical Activity

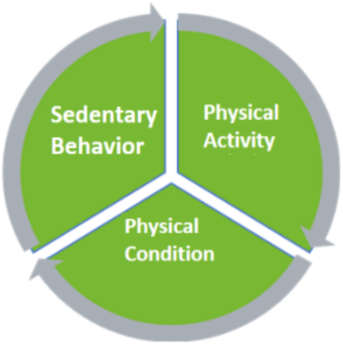


#### Endurance

| | Age Years | 1 VERY BAD | 2 BAD | 3 WEAK | 4 AVERAGE | 5 GOOD | 6 VERY GOOD | 7 EXCELLENT |
|---|---|---|---|---|---|---|---|---|
| Men | 20-24 | < 32 | 32-37 | 38-43 | 44-50 | 51-56 | 57-62 | > 62 |
| | 25-29 | < 31 | 31-35 | 36-42 | 43-48 | 49-53 | 54-59 | > 59 |
| | 30-34 | < 29 | 29-34 | 35-40 | 41-45 | 46-51 | 52-56 | > 56 |
| | 35-39 | < 28 | 28-32 | 33-38 | 39-43 | 44-48 | 49-54 | > 54 |
| | 40-44 | < 26 | 26-31 | 32-35 | 36-41 | 42-46 | 47-51 | > 51 |
| Women | 20-24 | < 27 | 27-31 | 32-36 | 37-41 | 42-46 | 47-51 | > 51 |
| | 25-29 | < 26 | 26-30 | 31-35 | 36-40 | 41-44 | 45-49 | > 49 |
| | 30-34 | < 25 | 25-29 | 30-33 | 34-37 | 38-42 | 43-46 | > 46 |
| | 35-39 | < 24 | 24-27 | 28-31 | 32-35 | 36-40 | 41-44 | > 44 |
| | 40-44 | < 22 | 22-25 | 26-29 | 30-33 | 34-37 | 38-41 | > 41 |

##### Cardiorespiratory fitness

- Based on repeated shuttle runs between two lines set apart
- Pace dictated by auditory beeps
- Adapted version: 2 × 10 m shuttle runs
- Maximal aerobic speed corresponds to the last completed stage before stopping
- $VO_2max$ (mL/kg/min) estimated using an equation
- Results compared to reference standards adjusted for sex and age

##### Solutions

Ex 1 : While walking, accelerate the pace over a short distance until slightly out of breath.

Ex 2 : Increase active transportation (e-bike or regular bicycle, non-motorized scooter, walking).

Ex 3 : Gradually increase the intensity and/or duration of physical activities.

Preventive Remediation for OptiMal StudentS

#### Appendix 11. Cardiac coherence exercises

**Session 1**

Group session: Students will perform an HRV-biofeedback exercise using emWave® Pro Plus Coherence Training software (v3.14.1, HeartMath Institute, Boulder Creek, CA, USA). During the first three minutes, the expert will explain the visual interface, including the pulse wave, instantaneous heart rate, and cardiac coherence score. Cardiac coherence is defined as a stable, synchronized pattern between heart rate variability (HRV), respiration, and autonomic nervous system activity. The expert will then present the breathing cursor that will guide the participants' inhalation and exhalation rhythm. From three to six minutes, participants will perform a paced breathing exercise at a rate of six breaths per minute. Between six and eight minutes, the participants will continue the paced breathing exercise while visualizing a breathing image that follows the rhythm—for example, they might imagine a sun or balloon expanding during inhalation and shrinking during exhalation. From eight to nine minutes, participants will close their eyes and remain silent for one minute, without any specific breathing instructions. This will provide data on participants cardiac coherence without further help from the experts. At nine minutes, the recording will be stopped.

Individual meeting: Students will be debriefed based on their cardiac coherence trace, with the expert explaining the different phases of the recording. During the first three minutes, the trace corresponds to the basal level. From three to six minutes, the student performed a three-minute breathing exercise; this phase will allow discussion of reactivity, peak coherence score, and stability. Between six and eight minutes, the student completed two minutes of paced breathing with a respiratory image. From eight to nine minutes, the student remained inactive for one minute, allowing the expert to evaluate how well the coherence score was maintained; this corresponds to the "fade-out speed."

**Session 2**

Group session: The students will perform an HRV-biofeedback exercise so that their resonant breathing frequency can be estimated using an adapted version of the Lehrer procedure (23–25). First, the expert will briefly remind participants of the visual interface during 3 minutes, followed by a one-minute period with eyes closed. They will students will engage in three minutes of paced breathing at a frequency of nine cycles per minute, followed by one-minute period with eyes closed. Next, students will perform three minutes of paced breathing at a frequency of ten cycles per minute, followed by a final one-minute period with eyes closed. The expert will identify the most effective breathing frequency to reach a high and sustainable cardiac coherence (this should be close to the student's breathing resonance). Finally, the students will be advised to practise at their most effective breathing frequency for further self-training (23–25).

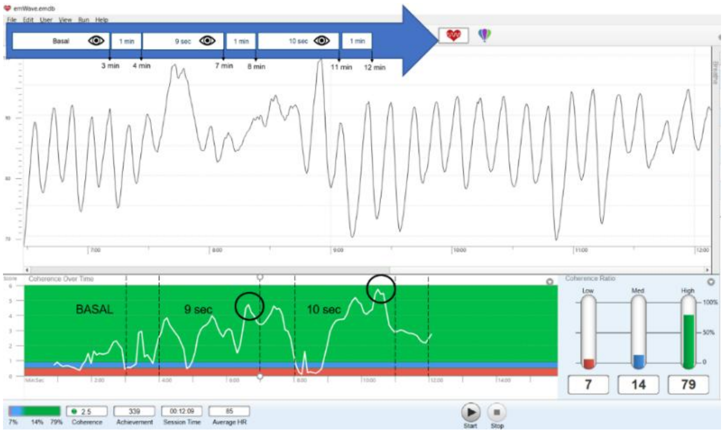

#### Appendix 12. Exercises to improve specific aspects of physical fitness

During the fifth session of the PROMESS-Group programme, students will receive individualized feedback on the results of the physical tests carried out before the intervention (T0) and described above. They will be given specific advice and exercises to improve their flexibility, power, muscle strength, and endurance. These exercises have been designed by experts in physical activity sciences and sports or by professionals with a degree in adapted physical activity; they are intended to be simple and accessible, and most of them can be done without specific equipment.

**Flexibility**
Objective: improve mobility in the hamstrings, back, and shoulders.
Reference tests: Sit-and-reach test and shoulder stretch test.
Recommended exercises:

- Exercise 1: Roll the shoulders forward and backward.
- Exercise 2: Bend the elbow, place the hand on the opposite shoulder blade, and gently pull the bent elbow toward the opposite side.
- Exercise 3: Raise the arms straight up for five seconds, then flex the spine forward and attempt to touch the feet.

**Power**
Objective: develop the ability to generate force quickly (working the anterior and posterior chains).
Reference test: Squat jump test.
Recommended exercises:

- Exercise 1: Take the stairs, perform bodyweight squats, and rise onto the toes 10 times.
- Exercise 2: Perform two-footed jumps onto mats of progressively increasing height.
- Exercise 3: In resistance training, use light loads with increased execution speed (e.g. squats, bench press, deadlift, forward/backward lunges).

**Strength**
Objective: to strengthen the major muscle groups, particularly the quadriceps, abdominals, and shoulder girdles.
Reference test: Maximal isometric quadriceps strength test.

- Exercise 1: Perform muscle-strengthening exercises (wall sit, core planks, abdominal exercises).
- Exercise 2: Perform 10 squats during an active break. Mountain climber exercise.
- Exercise 3: In resistance training, lift heavy loads for 10 repetitions (e.g. squats, bench press, deadlift, forward/backward lunges).

**Endurance**
Objective: improve cardiorespiratory capacity and recovery after exercise.
Reference test: 10-m shuttle run test.
Recommended exercises:

- Exercise 1: While walking, accelerate the pace over a short distance until slightly out of breath.
- Exercise 2: Increase active transportation (e-bike or regular bicycle, non-motorized scooter, walking).
- Exercise 3: Gradually increase the intensity and/or duration of physical activities.

#### Appendix 13. Assessment of student satisfaction with modules

Just before the end of each module, student satisfaction with the PROMESS-GROUP modules will be assessed using several 100-mm VASs. Each VAS will prompt the student to move a cursor to indicate their response. The cursor will start at the 0 position for each question. The answer cursor will range from 0 (absolutely not) to 100 (completely). These VASs were previously used during the PROMESS programme (19–21).

For each module, the student's satisfaction will be measured using a composite score, calculated as the mean of the specific score and the general score, described below.

**Specific stress satisfaction score.** Mean of the scores obtained for the "stress" VAS ("Do you think the intervention has helped you to lower your stress level?") and the "coping" VAS ("Do you think the intervention allowed you to better manage the stressful events you have encountered?").

**Specific sleep satisfaction score.** Mean of the scores obtained for the "sleep" VAS ("Do you think the intervention has helped you to improve your sleep?") and the "fatigue" VAS ("Do you think the intervention allowed you to decrease your fatigue?").

**Specific physical activity satisfaction score.** Mean of the scores obtained for the "sedentary behavior" VAS ("Do you think the intervention has helped you to decrease your sedentary behavior?"), the "physical activity" VAS ("Do you think the intervention has helped you to increase your physical activity level?"), and the "physical fitness" VAS ("Do you think the intervention has helped you to increase your physical fitness level?").

**General satisfaction score.** For the three modules, the general satisfaction score will be the mean of the score obtained for the "relevance" VAS ("Do you think the proposed goals were suitable for your daily life?") and the "sustainability" VAS ("Do you think you can sustain the performed changes on a daily basis?")

#### References


1. Andreacci JL, LeMura LM, Cohen SL, Urbansky EA, Chelland SA, Von Duvillard SP. The effects of frequency of encouragement on performance during maximal exercise testing. J Sports Sci. 2002;20(4):345-52.

2. Gouraud E, Connes P, Gauthier-Vasserot A, Faes C, Merazga S, Poutrel S, et al. Impact of a submaximal mono-articular exercise on the skeletal muscle function of patients with sickle cell disease. Eur J Appl Physiol. 2021;121(9):2459-70.

3. Mura M, Rivoire E, Dehina-Khenniche L, Weiss-Gayet M, Chazaud B, Faes C, et al. Effectiveness of an individualized home-based physical activity program in surgery-free non-endarterectomized asymptomatic stroke patients: a study protocol for the PACAPh interventional randomized trial. Trials. 2022;23(1):145.

4. Petersen NT, Taylor JL, Butler JE, Gandevia SC. Depression of Activity in the Corticospinal Pathway during Human Motor Behavior after Strong Voluntary Contractions. J Neurosci. 2003;23(22):7974-80.

5. Bachasson D, Villiot-Danger E, Verges S, Hayot M, Perez T, Chambellan A, et al. Mesure ambulatoire de la force maximale volontaire isométrique du quadriceps chez le patient BPCO. Rev Mal Respir. 2014;31(8):765-70.

6. Hamilton A, Balnave R, Adams R. Grip strength testing reliability. J Hand Ther Off J Am Soc Hand Ther. 1994;7(3):163-70.

7. Lagerström C, Nordgren B. Methods for measuring maximal isometric grip strength during short and sustained contractions, including intra-rater reliability. Ups J Med Sci. 1996;101(3):273-85.

8. Glatthorn JF, Gouge S, Nussbaumer S, Stauffacher S, Impellizzeri FM, Maffiuletti NA. Validity and reliability of Optojump photoelectric cells for estimating vertical jump height. J Strength Cond Res. 2011;25(2):556-60.

9. Mayorga-Vega D, Merino-Marban R, Viciana J. Criterion-Related Validity of Sit-and-Reach Tests for Estimating Hamstring and Lumbar Extensibility: a Meta-Analysis. J Sports Sci Med. 2014;13(1):1-14.

10. Holt LE, Pelham TW, Burke DG. Modifications to the Standard Sit-and-Reach Flexibility Protocol. J Athl Train. 1999;34(1):43-7.

11. Minick KI, Kiesel KB, Burton L, Taylor A, Plisky P, Butler RJ. Interrater reliability of the functional movement screen. J Strength Cond Res. 2010;24(2):479-86.

12. Gribble PA, Brigle J, Pietrosimone BG, Pfile KR, Webster KA. Intrarater reliability of the functional movement screen. J Strength Cond Res. 2013;27(4):978-81.

13. Cook G, Burton L, Hoogenboom B. Pre-Participation Screening: The Use of Fundamental Movements as an Assessment of Function – Part 1. North Am J Sports Phys Ther. 2006;1(2):62-72.

14. Léger LA, Lambert J. A maximal multistage 20-m shuttle run test to predict VO2 max. Eur J Appl Physiol. 1982;49(1):1-12.

15. Magee MK, White JB, Merrigan JJ, Jones MT. Does the Multistage 20-m Shuttle Run Test Accurately Predict VO2max in NCAA Division I Women Collegiate Field Hockey Athletes? Sports. 2021;9(6):75.

16. Cho HL, Park HY, Nam SS. Development of multistage 10-m shuttle run test for VO2max estimation in healthy adults. J Mens Health. 2022;18(1):1-8.

17. Torres G, Gordon NF, Constantinou D. Cardio-Respiratory Fitness and Cardiovascular Disease Risk Factors Among South African Medical Students. Am J Lifestyle Med. 2023;17(6):791-8.

18. Leger D, Esquirol Y, Gronfier C, Metlaine A. Republication de : Le travail posté et de nuit et ses conséquences sur la santé : état des lieux et recommandations. Médecine Sommeil. 2019;16(3):191-9.

19. Métais A, Omarjee M, Valero B, Gleich A, Mekki A, Henry A, et al. Determining the influence of an intervention of stress management on medical students' levels of psychophysiological stress: the protocol of the PROMESS-Stress clinical trial. BMC Med Educ. 2025;25:225.

20. Ruet A, Ndiki Mayi EF, Métais A, Valero B, Henry A, Duclos A, et al. Determining the influence of a sleep improvement intervention on medical students' sleep and fatigue: protocol of the PROMESS-Sleep clinical trial. BMC Med Educ. 2025;25:267.

21. Schlatter S, Mura M, Morel B, Loisel O, Aouidat M, Gouraud E, et al. Reducing Sedentary Behavior and Promoting Physical Activity in Medical Students: Protocol of the PROMESS-Physical Activity Clinical Trial [Internet]. Research Square; 2025 [cité 7 nov 2025]. Disponible sur: https://www.researchsquare.com/article/rs-7441836/v1

22. Métais A, Besnard L, Valero B, Henry A, Schott AM, Rode G, et al. Addressing medical students' health challenges: codesign and pilot testing of the Preventive Remediation for Optimal MEdical StudentS (PROMESS) program. BMC Med Educ. 2025;25:812.

23. Shaffer F, Meehan ZM. A Practical Guide to Resonance Frequency Assessment for Heart Rate Variability Biofeedback. Front Neurosci. 2020;14:570400.

24. Lehrer PM, Vaschillo E, Vaschillo B. Resonant Frequency Biofeedback Training to Increase Cardiac Variability: Rationale and Manual for Training. Appl Psychophysiol Biofeedback. 2000;25(3):177-91.

25. Lehrer P, Vaschillo B, Zucker T, Graves J, Katsamanis M, Aviles M, et al. Protocol for heart rate variability biofeedback training. Biofeedback. 2013;41(3):98-109.
